# Changes in serum CXCL13 levels are associated with outcomes of Colorectal Cancer Patients Undergoing First-Line Oxaliplatin-Based Treatment

**DOI:** 10.1101/2024.02.15.24302875

**Authors:** Sara Cabrero-de las Heras, Xavier Hernández-Yagüe, Andrea González, Ferran Losa, Gemma Soler, Cristina Bugés, Iosune Baraibar, Anna Esteve, Miguel Ángel Pardo-Cea, Anne Hansen Ree, Neus Martínez-Bosch, Maria Nieva, Eva Musulén, Sebastian Meltzer, Tania Lobato, Carla Vendrell-Ayats, Cristina Queralt, Pilar Navarro, Clara Montagut, Ferran Grau-Leal, David Camacho, Raquel Legido, Núria Mulet-Margalef, Eva Martínez-Balibrea

## Abstract

**Background:** Reliable biomarkers for precision medicine in metastatic colorectal cancer (mCRC) are needed. Blood biomarkers like chemokines may offer insights into overall tumor burden, yet, few prospective studies explore chemokine dynamics during treatment. This study investigates the behavior of a chemokine panel in mCRC patients during first-line oxaliplatin-based treatment, aiming to identify predictive and prognostic biomarkers.

**Methods:** Blood from oxaliplatin-treated mCRC patients was collected at three time points: before treatment (PRET), at response evaluation (EVAR), and at disease progression or last follow-up (LFUP). A custom 11-chemokine panel assessed serum chemokine levels by Luminex®, correlating them with treatment response, overall survival (OS), and progression-free survival (PFS) using the Cox proportional hazards models with the inverse probability weighting (IPW) approach. Additionally, immune system-associated gene expression was studied by Nanostring® in 15 primary tumor samples and correlated with CXCL13 expression, OS, and PFS. *In silico* analysis of 119 liver metastases from CRC patients, post neoadjuvant oxaliplatin-based treatment or untreated, evaluated CXCL13 expression’s correlation with immune cell infiltration, tertiary lymphoid structure (TLS) presence, OS, and PFS. Additionally, CXCL13 dynamics was studied by ELISA in 36 mCRC patients from the METIMMOX study control arm.

**Results:** Responders exhibited increased CXCL13 at EVAR, contrasting with non-responders whose levels decreased at EVAR and LFUP. Increased CXCL13 independently associated with improved PFS (median 14.5 vs. 8.9 months; HR = 0.34, p = 0.003) and OS (median 39.7 vs. 15.3 months; HR = 0.34, p = 0.003). CXCL13 expression correlated positively with an immunogenic tumor microenvironment, increased B cells, T cells (mainly CD8+) and enhanced OS. *In silico*, higher CXCL13 expression associated significantly with increased immune infiltration and improved OS. High CXCL13 expression was linked to the presence of TLSs, also associated with enhanced OS, especially in neoadjuvant-treated patients. Similar trends were obtained using the METIMMOX cohort.

**Conclusion:** The increase of CXCL13 levels in peripheral blood and its association with the formation of TLSs within the metastatic lesions, emerges as a potential biomarker indicative of the therapeutic efficacy in metastatic CRC patients undergoing oxaliplatin-based treatment.

## 1. Background

Colorectal cancer (CRC) ranks as the third most prevalent form of cancer globally, with 1.1 million new diagnostics annually, and is the second principal contributor to fatalities attributed to cancer^1^. The survival rate for localized instances stands at 70-90%, while for metastatic conditions it plummets to 10% at 5 years. New options like immunotherapy and target therapy have been included in the therapeutic arsenal of uncommon molecular CRC subgroups (ie dMMR-MSI-H, *BRAF* V600E or *HER* 2 amplified) in the recent years. However, the current standard of care for the vast majority of metastatic patients remains unchanged and it is based on chemotherapy (fluoropyrimidines, oxaliplatin and/or irinotecan), typically in conjunction with anti-EGFR (cetuximab or panitumumab; exclusively for patients with KRAS and NRAS wild-type) or anti-VEGF (bevacizumab and aflibercept) agents^2^. While these treatments do initially display efficacy, resistance often follows, leading to disease progression and death in the majority of patients. Unfortunately, we currently lack reliable biomarkers with adequate specificity and sensitivity for being implemented into our daily practice that would allow us to select the best therapeutic option for our patients and to track the mechanisms of resistance.

Several research groups, including ours, have investigated the potential role of certain chemokines as predictive biomarkers and contributors to cancer treatment resistance. Chemokines are small proteins secreted by cells that attract particular cells from the immune system. They bind to specific seven transmembrane G-protein-coupled receptors to execute their biological function. Different types of chemokines have varying characteristics, with CXC family chemokines being identified by the presence or absence of an ELR motif (Glu, Leu, Arg) situated upstream of the first conserved cysteine residue^3–5^. ELR+ CXC chemokines are chemotactic for neutrophils and angiogenic, whereas most ELR-CXC chemokines are angiostatic, attracting lymphocytes and natural killer (NK) cells. Chemokines play a vital role in the mediation of lymphoid tissue organogenesis, lymphocyte homing and hematopoiesis. However, they are also implicated in pathological processes such as cancer. In addition to their chemoattraction capabilities, chemokines have been associated with other functions including cell survival and proliferation, as well as the promotion or inhibition of angiogenesis^6,7^.

The available data indicates that tumor treatment could lead to the secretion of particular chemokines that modify the microenvironment of the tumor. Dr Joan Massagué and his team demonstrated that CXCL1 and CXCL2 overexpression in breast tumors fosters a microenvironment that promotes chemoresistance and concurrently primes tumor cells for survival in organs targeted for metastasis^8^. More recently, there have been reports indicating that chemotherapy-induced apoptosis can stimulate the release of chemotactic factors, specifically CXCL8, that entice neutrophils into the tumor. These neutrophils might interact with neighboring macrophages, leading to an immunologically adverse tumor microenvironment, ultimately resulting in tumor recurrence^9^. Our research demonstrates that the development of resistance to oxaliplatin in CRC cells leads to an upregulation of the Nuclear Factor kappa-light-chain-enhancer of activated B cells (NF-kB) transcription factor. This, in turn, results in increased secretion of CXCL1, CXCL2 and CXCL8. Partial reversion of the resistant phenotype was observed by blocking either the NF-kB pathway or chemokine transcription^10^. All of the aforementioned factors, coupled with the fact that chemokines are secreted and detectable in the peripheral blood of patients, have positioned them as a focal point of cancer biomarker research. Nonetheless, there is a scarcity of studies that have systematically assessed the potential function of CXC chemokines as biomarkers of response to treatment in CRC. The aim of this study was to objectively assess the secretion dynamics of 11 CXC chemokines in the peripheral blood of individuals with metastatic CRC undergoing oxaliplatin-based regimens, using the Luminex technique. The data was analyzed to establish any correlation between the patients’ clinical outcomes and prognosis. Of all the chemokines analyzed, our findings suggest that the rise in CXCL13 following 12 treatment cycles is a positive predictive factor potentially linked to the enhancement of an immunogenic microenvironment and the formation of tertiary lymphoid structures (TLS).

## 2. Patients and methods

### 2.1. Study design and patients

This study was prospective and longitudinal, conducted at five different hospitals in Catalonia, Spain: Dr. Josep Trueta Hospital (Girona), Vall d’Hebron Institute of Oncology (VHIO, Barcelona), Moises Broggi Hospital (Sant Joan Despí), Duran I Reynals Hospital (L’Hospitalet de Llobregat), and the coordinating center, Germans Trias i Pujol University Hospital (Badalona). The study received approval from the Ethics Committee of Germans Trias I Pujol University Hospital under the internal reference number IP-16-115. Patients included in the study had either colon or rectal adenocarcinoma confirmed through histological assessment, with measurable metastatic disease (Response Evaluation Criteria in Solid Tumors [RECIST]; Version 1.1). Patients were aged 18 or older, had an Eastern Cooperative Oncology Group performance status (PS) of 0-2, and adequate renal function. All patients were treated with oxaliplatin-based regimens, including: FOLFOX, CAPOX, FOLFOX or CAPOX combined with Bevacizumab, FOLFOX with Panitumumab (only for RAS wild-type patients), and FOLFOX or CAPOX with Cetuximab (only for RAS wild-type patients). All detailed regimens are shown in Supplementary Table S1.

All patients received proper information and signed the corresponding informed consent. Patient inclusion, follow-up, and sample collection were executed for a period of 52 months (November 2016 to April 2021) and were partially impacted by the COVID-19 pandemic.

### 2.2. Sample collection, processing and storage

Serum samples were collected at various time points throughout the disease (Figure 1). The initial sample (PRET) was taken during a routine blood test prior to commencing treatment. The second sample (EVAR) was collected during the response evaluation, typically 3 months (12 weeks) after the PRET sample was obtained. Finally, the third sample (LFUP) was procured at the time of tumor progression, the last follow-up or the end of the study, whichever came first. For patients with progressive disease at the first response evaluation, only two samples were available (PRET and LFUP). Additionally, when possible, a primary tumor sample fixed in formalin and embedded in paraffin (FFPE) was gathered. One SST™ tube (BD Vacutiner®, BD) containing 5mL of whole blood was collected per patient and time-point. The extraction, collection, and storage of samples followed a consistent process in all hospitals. After the SST was extracted, it was centrifuged at 1000g for 15 minutes at 4°C. Following that, the serum was divided into six 500μL aliquots and placed in Matrix™ 2D barcoded clear polypropylene opentop storage tubes by Thermo Scientific. This process was carried out in a biological safety cabinet and then stored in a Matrix 96-format 2D barcoded storage microplate, also by Thermo Scientific. Aliquots in the microplates were stored in an ultra-low temperature freezer (-80°C) at the IGTP Biobank to guarantee the correct storage and preservation of serums until their analysis.

**Figure 1.**
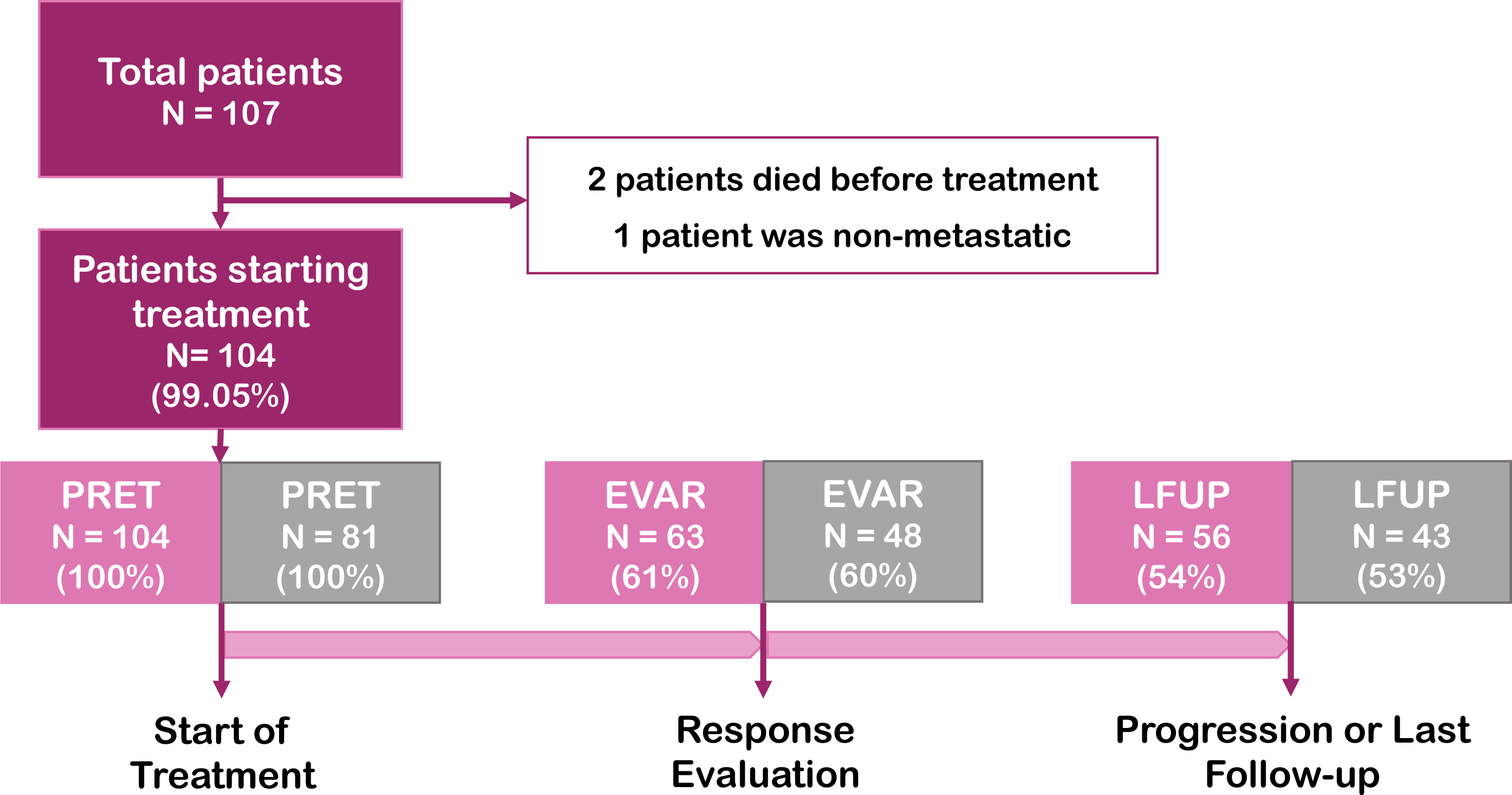
Study design. Study design and number of cases included in total and by type of sample collected (PRET, EVAR, LFUP). A distinction is made between total cases (pink) and cases that did not receive radical surgery during the study (grey).

### 2.3. Luminex® analysis

To determine the levels of CXC chemokines in all samples, we employed a custom panel named Bio-Plex Pro™ Human Chemokine multiplex Assay (Bio-Plex Pro™ Human Chemokine Panel, 171-AK99MR2, Bio-Rad). This specific test had pre-coated CXCL1, CXCL2, CXCL5, CXCL6, CXCL8, CXCL9, CXCL10, CXCL11, CXCL12, CXCL13, and CXCL16 antibodies that facilitated the detection of all chemokines in one well of a 96-well plate. The entire assay was completed within 4-5 hours. The lyophilized mixture of standard analytes was reconstituted, and serum samples were thawed before undergoing serial dilutions using the manufacturer’s guidelines. Each sample was pre-diluted with PBS 10 times (1:10), and two replicates were prepared. All subsequent steps were carried out in accordance with the manufacturer’s instructions, and readouts were performed using a Luminex® 200 instrument at the IGTP Flow Cytometry core facility. The standard curve was created via a five-parametric logistic regression non-linear model by utilizing the Xponent 3.1 software (Luminex Inc.). After ensuring cross-reactivity across all analytes, markers were categorized and grouped based on the dilution factor. To determine the concentration of each metabolite (pg/mL) in the serum, measurement values were interpolated. During the experiment, intra-assay precision ranged from 2% to 4%, and inter-assay precision ranged from 2% to 8%. To mitigate variability, we conducted two daily assays for three consecutive days.

### 2.4. CXCL13 *in silico* analysis

An *in silico* analysis was conducted using the existing Affymetrix Human Transcriptome 2.0 data from CRC liver metastases, as detailed in ^11,12^. Gene expression, clinicopathological annotations, and data pertaining to patient survival were obtained from the GSE159216 accession number at the GEO database. Raw intensity CEL files underwent background correction, normalization, gene-level summarization, and log2 transformation through the reliable multi-array average (RMA) technique, performed within the justRMA function of the affy R package (v1.80)^13^, as well as the custom Entrez CDF file (v22) from Brainarray (http://brainarray.mbni.med.umich.edu). HGNC gene symbols were obtained by converting Entrez IDs utilizing the biomaRt package (v2.52.0). Specifically, we analyzed data collected from 119 patients with CRC, consisting of 48 female and 71 male individuals. Among them, 15 patients had not undergone any prior treatment before hepatic resection, whereas 104 patients had received neoadjuvant treatment based on OXA. Out of the entire cohort, 50 patients had KRAS mutations, 7 had NRAS mutations, and 62 had no mutations in either of these genes. To determine the presence or absence of TLSs in patients of this dataset, we utilized three previously described genetic signatures: the 12-Chemokine^13^, the TFH cell^14^, and TH1 cell and B cell signatures^15^. Gene set scores were calculated through single sample Gene Set Enrichment Analysis (ssGSEA) utilizing the R package GSVA (v1.44.5)^16^. Normalized gene expression was utilized to determine the abundance of immune cell populations by using the MCP-counter algorithm^17^.

### 2.5. Nanostring experiments

RNA from FFPE CRC primary tumor tissue samples was extracted with the truXTRAC FFPE RNA Kit (Covaris), using the Covaris Adaptive Focused Acoustics and following manufacturers’ instructions. A final RNA yield of 150ng as well as an A260/A280 ratio of 1.7–2.3 and an A260/A230 ratio of 1.8–2.3 per sample, is recommended for subsequent analysis with the NanoString nCounter® Elements Technology. RNA quality was evaluated using an Agilent Bioanalyzer TapeStation system (Agilent, IGTP Genomics Facility). A customized panel by NanoString technology nCounter interrogating 21 immune-related genes and associated controls was used (Supplementary Table S2).

Normalization was performed using nSolver Analysis Software 4.0. Geometric mean was used to compute normalization factor into a two steps process, positive control normalization and codeset content normalization with ACTB, HPRT1 and TUB as reference genes. Low quality samples were removed during raw data import and normalization quality control processes. Normalized expression data was log2 transformed for further analysis.

### 2.6. Validation cohort

As an independent cohort to validate our results, we used samples from patients enrolled in the randomized, controlled, phase II trial METIMMOX (NCT03388190). Briefly, 80 patients were randomly assigned to receive either eight cycles of the oxaliplatin-based Nordic FLOX regimen Q2W (oxaliplatin 85mg/m2 on day 1 and bolus 5-fluorouracil 500mg/m2 and folinic acid 100 mg on days 1 and 2; control arm) or two cycles of FLOX Q2W before two cycles of nivolumab Q2W (240 mg flat dose) in a repeat sequential schedule to a total of eight cycles (experimental study arm). After eight treatment cycles, all patients proceeded to a treatment break, with CT evaluations every 8 weeks until disease progression, whereupon treatment was restarted. Patients were treated until the first confirmed disease progression on active therapy, intolerable toxicity, withdrawal of consent or death, whichever came first. A detailed description of the study has been published previously^18^.Only patients included in the control arm of the study were used in this project. Consecutive serum samples were collected at baseline (N = 36) and after 4 (N = 33), 8 (N = 28), 12 (N = 23) and 16 weeks (N = 17). Last sample was obtained at end of treatment (N = 16). Nineteen patients withdrew the study due to either severe toxicity or disease progression within the first 16 weeks of treatment, contributing to a falling number of serum samples available for analysis. Patients’ characteristics are shown in Supplementary Table S3. Whole blood samples were collected in Vacuette® serum tubes, left at room temperature for at least 30min and centrifuged at 2200G for 10min. Further serum was aliquoted into 1mL vials and frozen and stored at - 80°C until CXCL13 ELISA analysis.

### 2.7. ELISA assay

To evaluate the CXCL13 levels in samples from the validation cohort we performed an ELISA assay using the Human CXCL13/BLC/BCA-1 Immunoassay Quantikine ELISA kit (R&D Systems). Serum samples were diluted 1:2 using the calibrator diluent (RD6-41) of the kit. Human CXCL13 standard was reconstituted in distilled water to further prepare serial dilutions as per the manufacturer’s instructions. Optical density (OD) was measured at 450nm and corrected at 540nm. The median value of two technical replicates was used for further analyses. The standard curve was generated by linearizing the OD values to the known human CXCL13 concentrations from the standards. Samples OD measurements were multiplied by the dilution factor (x2) and interpolated in the standard curve to obtain CXCL13 (pg/mL) concentration in the serum.

### 2.8. Statistical methods

#### Analysis of Luminex Study Findings

Patient characteristics were described using frequencies for categorical variables and the median with interquartile range (IQR) for continuous variables. The CXCs levels were analyzed at defined time-points using the paired Wilcoxon signed rank test. The Kruskal-Wallis test was used to assess the association between clinical and demographic factors and chemokine values at each time-point. To calculate the overall survival (OS), follow-up time (in months) was determined from the day of initial treatment commencement until death from any cause or latest follow-up, whichever took place first. Progression-free survival (PFS) follow-up time (in months) was measured from the day of initial treatment commencement until progression, death from any cause or latest follow-up, whichever took place first. For patients who had surgery after their first treatment, the follow-up time for PFS was stopped at the date of surgery. We estimated the median OS and median PFS using the Kaplan-Meier (KM) method and compared survival curves stratified by patient characteristics with the log-rank test. The associations between chemokines’ dynamics, defined as increase or decrease levels from PRET to EVAR, and the risk of death (or progression/death) were analyzed using Cox proportional hazards models with the inverse probability weighting (IPW) approach which allows us to account for confounding bias. The probability of a patient having an increase or decrease in chemokines’ levels was estimated with logistic regression models considering PS, tumor location, number of metastatic sites and radical surgery as common regressors for the exposure status and the outcomes. Stabilized weights were computed from estimated probabilities and used in the Cox regression models to reduce for imbalance in measured confounders between increase or decrease exposure groups. Unadjusted Hazard ratios (HRs) and IPW-adjusted HRs with their 95% confidence intervals (CIs) were reported. The levels of CXCs were transformed to log2 and a heatmap with hierarchical clustering was generated using the Euclidean distance. All statistical analyses were conducted using the statistical software R v.4.1.2.

#### CXCL13 *in silico* analysis

Pearson’s correlation coefficients were computed using the cor and cor.test functions from the stats R package (v4.3.1). The ggsurvplot function from the survminer R package (v0.4.9) was used to construct Kaplan-Meier plots. The features were partitioned into two groups determined by their median values. Furthermore, multivariate Cox regression analysis was executed to obtain hazard risks and confidence intervals by including gender as a covariate, employing the coxph function from the coxph R package (v3.5-7). The wilcox.test function from the R package stats v4.3.1 was used to perform a two-tailed Wilcoxon Rank-Sum Test.

#### Nanostring experiments

We calculated Spearman correlation coefficients using the cor and cor.test functions from the stats R package (v4.3.1). We constructed Kaplan-Meier plots which include the log-rank test p-value, using the ggsurvplot function from the survminer R package (v0.4.9). We divided the features into two groups, using median values.

#### Validation studies

We analysed the data using IBM SPSS Statistics for Mac version 28 or GraphPad Prism v9.5.0. Data is presented either as a median and interquartile range or as the number of events and percentage of the total. Linear regression was calculated using Pearson’s r-test, correlations were determined using Spearman’s rho test, and differences between groups were analysed using the Kruskal-Wallis test or Mann-Whitney U test where appropriate. Survival disparities were evaluated through the log-rank test and exhibited utilizing Kaplan-Meier curves, or Cox proportional hazard models were conducted utilising gender as a covariate and exhibited as the hazard ratio (HR) and 95% confidence intervals (95% CI). All analyses were two-sided, and P values of less than 0.05 were deemed statistically significant.

## 3. Results

Of the 107 patients enrolled in the study, two died before treatment and one had non-metastatic disease. Therefore, serum samples were collected from 104 patients prior to the first treatment cycle (PRET). Of these, 63 samples were obtained at the time of response evaluation (EVAR) whereas 56 samples were collected at the time of disease progression (N = 47) or last follow-up (N = 9) (LFUP). Similar proportions were observed for patients who did not undergo radical surgery (Figure 1). Sixty four percent of patients demonstrated positive response to first-line treatment, and only 11.5% exhibited progressive disease. The median progression-free survival (PFS) and overall survival (OS) were 11.01 (95% CI 9.8 – 13.1) and 25.32 (95% CI 19.6 – 36.0) months, respectively. All patients’ characteristics are shown in Table 1.

**Table 1.** Patients’ characteristics.

| **VARIABLES** | **ALL N = 104** |
| --- | --- |
|  | **N (%)** |
| **Age at Diagnosis [IQR]** | 66.5<br>[58.8;73.0] |
| **Sex** |  |
| Male | 74 (71.2%) |
| Female | 30 (28.8%) |
| **Performance Status** |  |
| 0 | 41 (39.4%) |
| 1 | 59 (56.7%) |
| 2 | 4 (3.8%) |
| **Primary Tumor Site** |  |
| Colon | 66 (63.5%) |
| Rectum | 38 (36.5%) |
| **Metastatic Site** |  |
| Liver | 51 (49.0%) |
| Lung | 11 (10.6%) |
| Liver + Lung | 31 (29.8%) |
| Other | 11 (10.6%) |
| **Number of metastasis** |  |
| < 5 Metastasis | 32 (30.8%) |
| ≥ 5 Metastasis | 72 (69.2%) |
| **First Line Treatment** |  |
| FOLFOX | 33 (31.7%) |
| CAPOX | 4 (3.9%) |
| FOLFOX-Bevacizumab | 38 (36.6%) |
| FOLFOX-Cetuximab | 9 (8.7%) |
| CAPOX-Bevacizumab | 2 (1.9%) |
| CAPOX-Cetuximab | 1 (1%) |
| FOLFOX-Panitumumab | 17 (16.3%) |
| **Response Evaluation** |  |
| Complete Response (CR) | 1 (1%) |
| Partial Response (PR) | 67 (64.4%) |
| Stable Disease (SD) | 24 (23.1%) |
| Progressive Disease (PD) | 12 (11.5%) |
| **Second Line** |  |
| Yes | 60 (47.7%) |
| No | 44 (42.3%) |
| **Surgery During Study** |  |
| No Surgery | 71 (68.3%) |
| Only Primary Tumor | 10 (9.6%) |
| Only Metastasis | 9 (8.7%) |
| Primary and Metastasis | 14 (13.4%) |
| **Resected Metastatic Organs** |  |
| Liver | 18 (78.3%) |
| Lung | 2 (8.7%) |
| Peritoneum | 3 (13%) |
| **Microsatellite instability** |  |
| MSI | 5 (4.8%) |
| MSS | 75 (72.1%) |
| Unknown | 24 (23.1%) |
| **BRAF and RAS mutation** |  |
| BRAF mut | 4 (3.8%) |
| NRAS mut | 9 (8.7%) |
| KRAS mut | 46 (44.2%) |
| WT | 43 (41.3%) |
| Unknown | 2 (1.9%) |
| **Immune System Known Alterations** |  |
| Autoimmune Disease | 2 (1.9%) |
| Patient with a Transplant | 1 (1.0%) |
| Major Surgery Last Month | 3 (2.9%) |
| Unknown | 98 (94.2%) |
| **Exitus** |  |
| Yes | 60 (57.7%) |
| No | 44 (42.3%) |

Patients with a PS of 1 or greater, or those who did not respond to treatment according to RECIST v1.1 criteria, faced a substantially higher risk of progression and death. Furthermore, patients with multiple metastases (5 or more) had a poorer OS as compared to those with less than 5. Significantly, 23 cases (22.1%) underwent radical surgery, either for metastasis alone or both the primary tumor and metastasis, at some point following response evaluation during the study. As anticipated, this led to a notably improved OS and PFS in the patients who underwent surgery (Supplementary Table S4). Considering that the presence or absence of the primary tumor and/or metastases could impact the secretion of chemokines, this variable, along with the others that affected OS and/or PFS were subsequently accounted for in multivariate Cox regression models (see material and methods section). Notably, our study collected data on potential immune system alterations. Specifically, two patients had an autoimmune disease, one patient received a transplant, and three patients underwent major surgery within the last month prior to enrollment. In these patients, baseline levels of all studied chemokines did not differ from the rest of the cohort (data not shown) and they were therefore included in the study.

The Luminex technique allowed for the detection of all examined chemokines, yet their distributions exhibited non-normal curves. Figure 2A shows the correlations between the different chemokines studied at the basal level; of note, CXCL1 and CXCL6, or CXCL9 and CXCL10, showed a statistically significant correlation and a “r” value higher than 0.7. Interestingly, CXCL16 showed the lowest correlation values with all chemokines. In PRET samples, chemokines were distributed into two distinct clusters based on their higher (CXCL2, CXCL5, CXCL12, CXCL16, CXCL1 and CXCL10) or lower basal concentrations (CXCL9, CXCL8, CXCL6, CXCL11 and CXCl13) (Figure 2B). It is noteworthy that most of the chemokines categorized as pro-tumoral or pro-angiogenic were clustered together, as were those classified as anti-tumor or anti-angiogenic. Multiple associations were discovered between chemokine levels and clinicopathological features, particularly in baseline samples. It is of note that CXCL1, CXCL5, and CXCL8 PRET levels were markedly higher in patients with liver and lung metastases compared to other metastatic sites (Figure 3A). Moreover, baseline levels of CXCL1, CXCL2, CXCL6, and CXCL8 were significantly higher in patients with multiple metastases compared to those with oligo-metastatic disease (Figure 3B). We performed comparisons with all the categorical variables listed in Table 1; however, only those in which we found any statistically significant associations can be found in Supplementary Figure S1.

**Figure 2.**
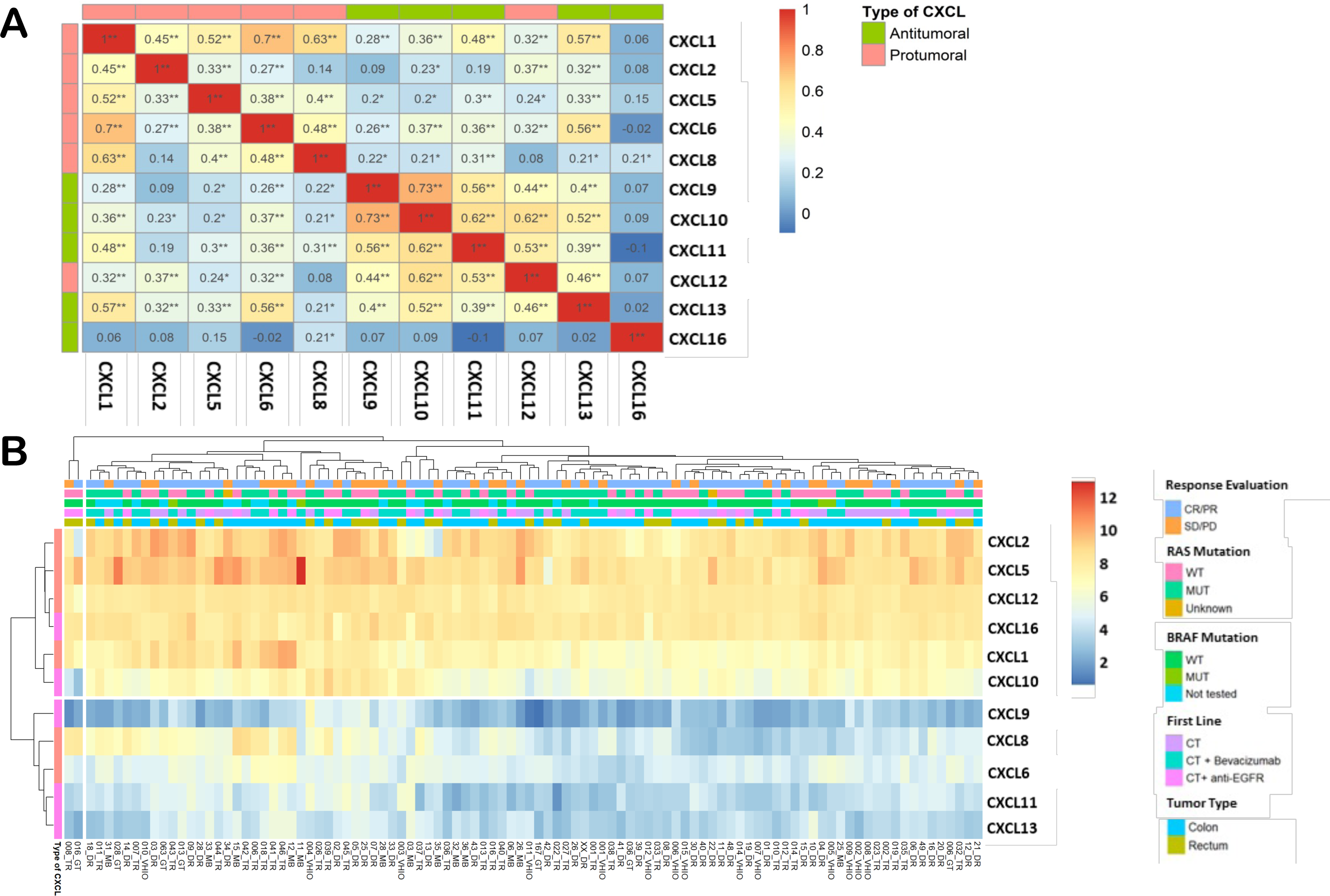
Baseline distribution of the examined chemokines. (A) Correlation table illustrating the interrelationship among diverse chemokines based on concentration values in the PRET sample. (B) Heat map presenting unsupervised clustering of the studied chemokines, reflecting their concentration values in the PRET samples.

**Figure 3.**
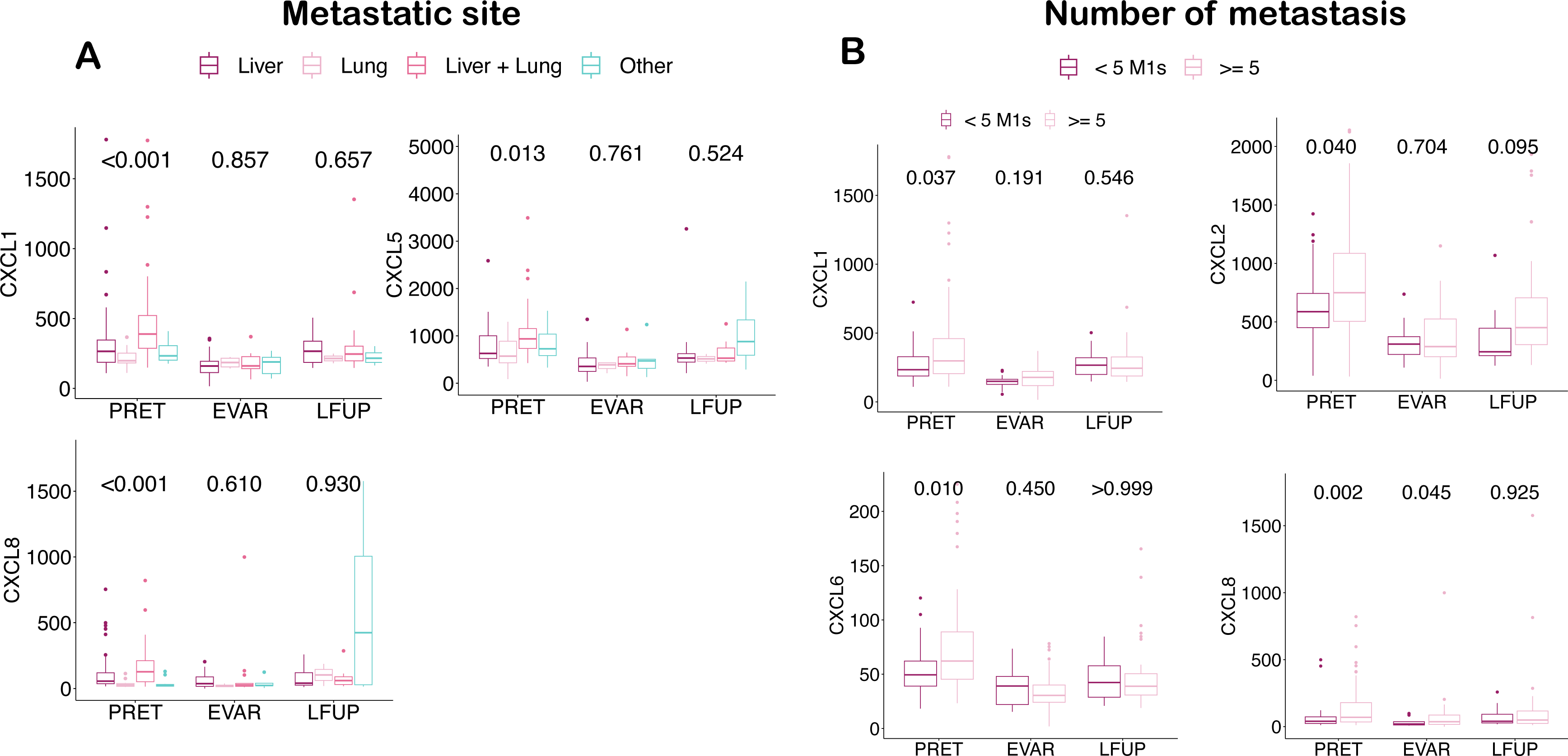
Association of select chemokines with the number and site of metastases. (A) Box plots presenting the median concentration (horizontal line) and standard deviation (SD, vertical line) of the specified chemokines in each study sample, categorized by the site of metastasis. (B) Box plots displaying the median concentration and SD of the specified chemokines in each study sample, stratified by the number of metastases. The p-values resulting from Kruskal-Wallis test are provided at the top of each graph.

We analyzed the dynamics of the change in the mean chemokine concentrations according to patients’ response to treatment. Pro-tumoral chemokines generally decreased in the EVAR sample compared to PRET and increased in the LFUP sample compared to EVAR, with the exception of CXCL8 which increased in both cases. However, when we categorized the patients as responders and non-responders, we can see that in the former group, CXCL8 remained relatively stable, while in the latter group, it shows a significant increase, particularly in the LFUP sample (Figure 4A). As mentioned before, it is important to note that the LFUP samples mostly correspond to the time of disease progression, but in some cases, the patients had not yet progressed. Thus, we conducted a similar analysis by excluding patients who had not yet progressed (N = 9) at the time of LFUP sample collection. However, we did not observe any difference in the pattern of change (Figure 4B).

**Figure 4.**
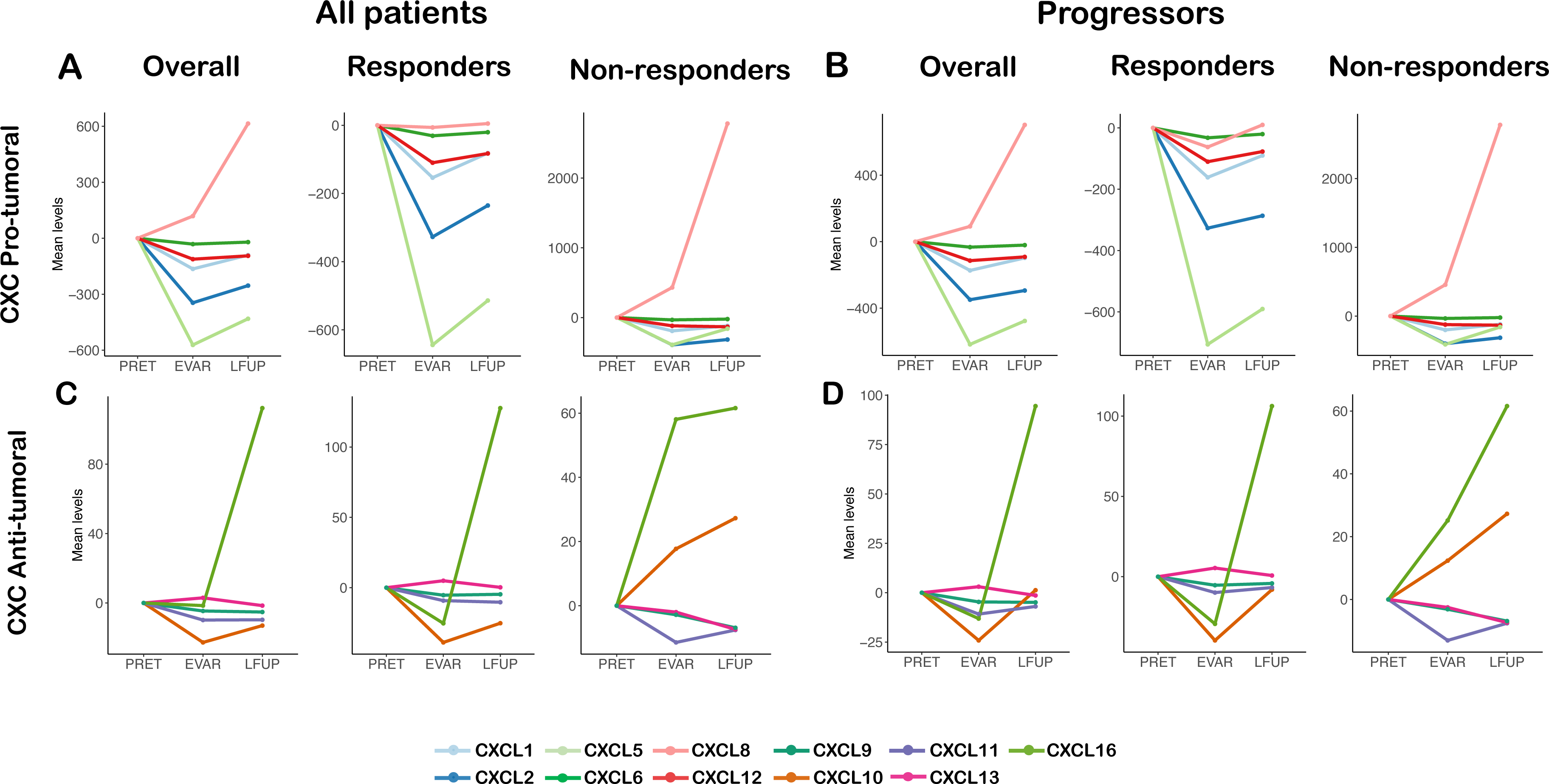
Dynamics of CXC chemokines. The figure depicts the mean concentration values of all examined chemokines in each sample, with the PRET sample serving as the reference value. (A) Pro-tumor chemokines across the entire cohort and in relation to treatment response. (B) Pro-tumor chemokines in relation to treatment response excluding cases without progression at the time of LFUP sample collection (N = 9). (C and D) Analogous results in anti-tumor chemokines.

The dynamics of change for anti-tumor chemokines were less homogeneous, with some following a V-shaped pattern and others following an inverted V-shaped pattern (Figure 4C). Notably, CXCL16 and CXCL13 exhibited unique behavior. In responders, CXCL16 decreased in EVAR samples but increased greatly in LFUP ones. In non-responders, this increase already occurred in the EVAR sample. On the other hand, CXCL13 increased in the EVAR and decreased in the LFUP in responders while in non-responders, the chemokine progressively decreased in both EVAR and LFUP. No significant differences were observed when patients who did not progress at the time of LFUP sample collection were excluded (Figure 4D).

Only changes in levels of CXCL13 at the EVAR time-point in comparison with the baseline were significantly linked with OS and PFS in both univariate and multivariate analyses, among all the chemokines examined (Supplementary Table S5 and Figure 5A). More precisely, patients who exhibited an increase in CXCL13 at the EVAR sample in relation to the baseline sample had a reduced risk of death (median OS of 39.7 months for patients with increase values compared to a median OS of 15.3 months for patients with decrease values; HR = 0.34; 95% CI: 0.16 - 0.69; p = 0.003) and disease progression (median PFS of 14.5 months in patients with increase values compared to a median PFS of 8.9 months in patients with decrease values; HR = 0.34; 95% CI 0.17 - 0.69; p = 0.003). These findings were further confirmed through analysis of patients who did not undergo radical surgery (median OS of 29.4 months in patients with increase values compared to a median OS of 14.1 months in patients with decrease values; HR = 0.32; 95% CI 0.15 – 0.68; p = 0.003; patients with increase values had a median PFS of 14.3 months compared to 8.2 months for patients with decrease values; HR = 0.31; 95% CI 0.15 - 0.62; p = 0.001) (Figure 5B). Then, we explored the dynamics from the EVAR time-point onwards and we observed that in those patients where CXCL13 increased in the EVAR sample, a mean decrease was observed in the LFUP values. In contrast, in the group in which CXCL13 decreased in the EVAR, the mean levels of this chemokine remained similar in the LFUP sample (Supplementary Figure S2A). In addition, this analysis was repeated excluding the cases that did not progress, obtaining very similar results (Supplementary Figure S2B)

**Figure 5.**
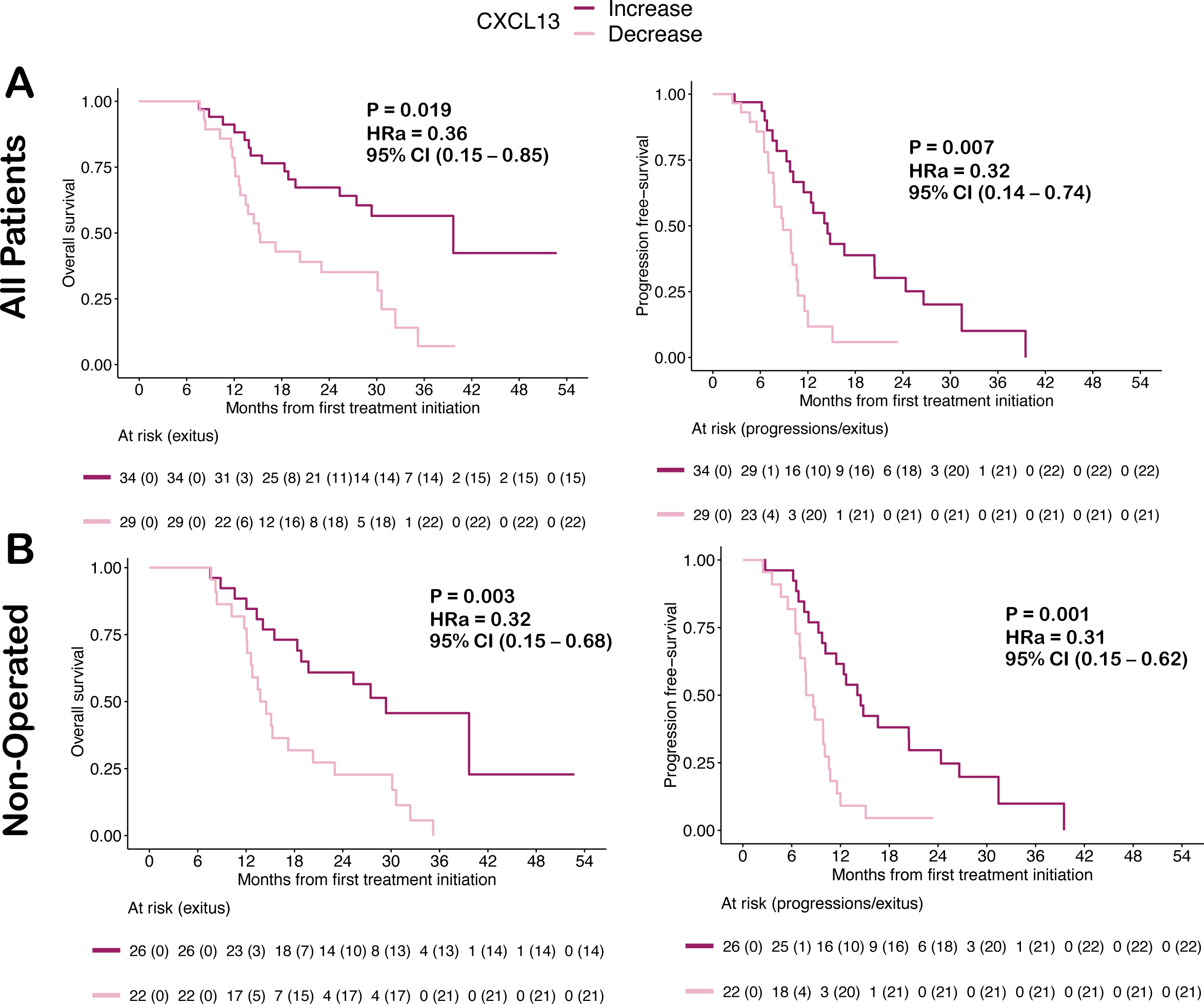
CXCL13 dynamics and correlation with OS and PFS. The Kaplan-Meyer plots show the OS (A) and PFS (B) curves of patients categorized according to the increase or decrease of CXCL13 in the EVAR sample with respect to PRET. Results are shown for the entire cohort and for patients who did not undergo radical surgery during the study, as indicated. The p-values, adjusted hazard ratios (HRa) and 95% confidence intervals (CI) shown were obtained from the multivariate COX regression analysis.

CXCL13 was initially recognized as a B-cell chemoattractant that is vital for the formation of lymphoid tissues. The CXCL13:CXCR5 axis significantly influences cellular interactions that govern lymphocyte infiltration into the tumor microenvironment, which in turn can influence the efficacy of cytotoxic drugs and immunotherapies^19,20^. Therefore, our aim was to examine the correlation between CXCL13 and tumor immunogenicity in our patients. Primary tumors from 26 patients were collected and used for a gene expression study via Nanostring® technology.

The panel of selected genes (Supplementary Table S2) aimed to identify populations of Dendritic Cells (DCs), Cytotoxic T cells, CD8+ T cells, Natural Killer cells (NKs), B cells, T-Follicular Helper Cells (THF), Regulatory T cells (Tregs), as well as CXCL13 expression^19,21,22^. After undergoing RNA quality and normalization filters, a total of 15 samples were eligible for analysis. It was observed that samples exhibiting high *CXCL13* expression generally displayed elevated expression levels mainly for genes associated with B cell and CD8 T cell populations (Figure 6A); indeed, *CXCL13* expression was found to have a statistically significant positive correlation with *CD79A*, *CD19*, *HLADOB*, and *CD40* (B cells) as well as *CD8A* and *CD8B* (CD8 T cells) (Figure 6B). There was also a trend towards a positive correlation with the genes *MS4A1* (B cells), *CTRAM* (CD8), *GZMM* (NKs) and *GZMK* (cytotoxic T cells). There was also a weak positive correlation, although not significant, with *CXCR5* and *IL21* (TFH) (Figure 6B and Supplementary Figure S3). Patients with high *CXCL13* expression displayed better OS and a trend to a better PFS as compared to those with low *CXCL13* expression (p-value for log-rank test 0.027 and 0.100, respectively). Additionally, high *CD79A* expression (B cells) was also linked to a better prognosis among all studied genes, with outcomes similar to those observed for CXCL13 (Figure 6C).

**Figure 6.**
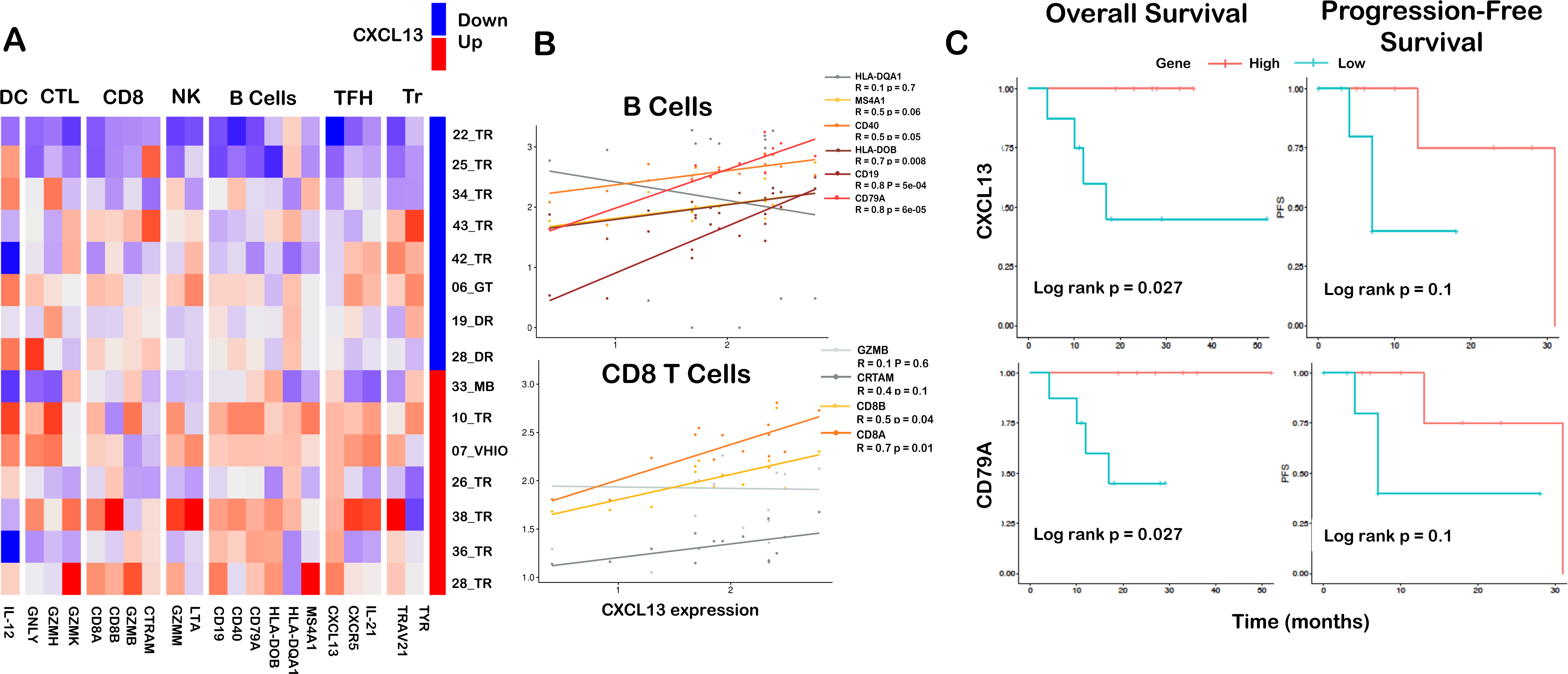
CXCL13 and Immune Tumor Microenvironment. (A) Heat map displaying the gene expression profile analyzed using Nanostring® in the primary tumors of study patients. The analyzed genes are presented at the bottom of the figure, grouped based on the immune cell populations they represent (depicted at the top). Samples are supervisedly clustered according to CXCL13 expression values above (red) or below (blue) the median expression level. Case numbers are shown on the right side of the heat map. (B) Scatter plots illustrating the correlation between CXCL13 expression and representative genes of B lymphocytes and CD8+ T cells, as indicated. The legend provides p and r values corresponding to the Spearman test. (C) Kaplan-Meier curves showing OS and PFS for patients categorized based on tumor expression of CXCL13 and CD79A above (high) or below (low) the median, as indicated.

In light of these results, we sought to confirm and replicate them by analyzing transcriptomic data sourced from liver metastases from a patient cohort that predominantly underwent oxaliplatin-based neoadjuvant regimens. To this end, we used data sourced from GSE159216 (see materials and methods). First, using the MCP counter algorithm, we assessed correlations of *CXCL13* gene expression with infiltrating cells in the tumor microenvironment, such as immune cells, fibroblasts and endothelial cells (Figure 7A). We observed that *CXCL13* expression correlated positively with all cell populations, with the most robust associations (correlation coefficients greater than 0.4) with B and cytotoxic cells. When focusing solely on treated patients, the correlations with B cells and cytotoxic cells demonstrated the greatest strength, and the correlation with T cells also showed significant results with a correlation coefficient of 0.4. In untreated (naive) patients, the number of statistically significant correlations was notably decreased. Secondly, we analyzed the association of *CXCL13* expression with survival, observing that increasing *CXCL13* expression was associated with improved OS among treated patients. However, when considering all patients or only those who did not receive treatment, the correlation between *CXCL13* gene expression and OS was not found to be statistically significant (figure 7B).

**Figure 7.**
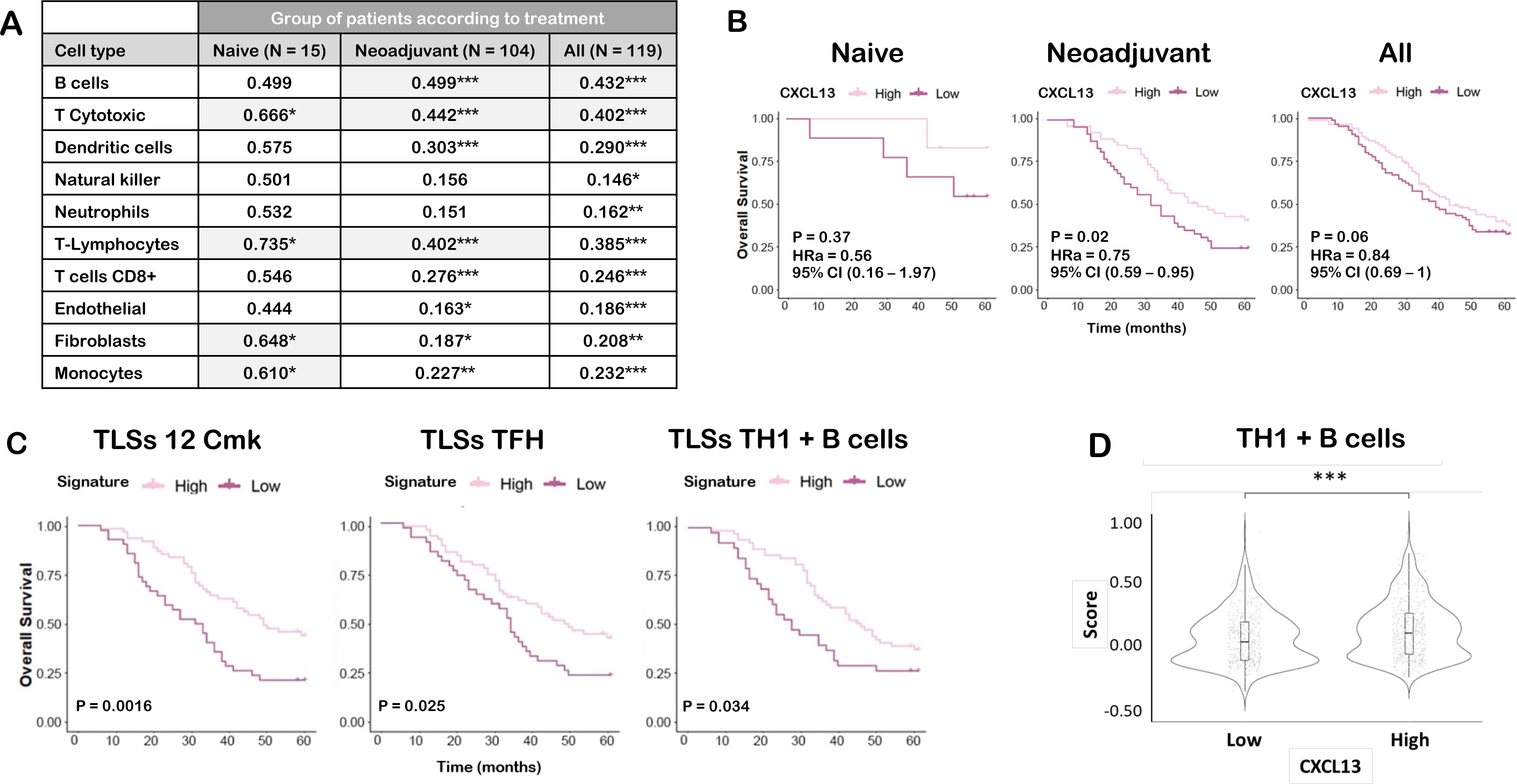
*In silico* Study of the association between CXCL3 and Presence of Tertiary Lymphoid Structures (TLSs). (A) MCP counter analysis investigating the correlation between CXCL13 expression and specified cell populations in the GSE159216 cohort. The displayed R and p values (*) resulted from Pearson’s analysis. Statistically significant values with R > 0.4 are shaded in light gray. (B) Kaplan-Meyer plots depicting OS for patients in the same cohort, categorized by CXCL13 expression above (High) or below (Low) the median, and stratified by the indicated treatment conditions. Corresponding p-values and HRa were derived from COX regression analysis. (C) Kaplan-Meyer curves illustrating OS for patients who received neoadjuvant treatment, categorized based on expression above (High) or below (Low) the median of gene signatures associated with TLSs. The p-values correspond to the Log-Rank test. (D) Violin plots presenting the correlation of CXCL13 expression with the gene signature of TLSs TH1 + B cells. The p value (*) corresponds to the Wilcoxon Rank-Sum test. *** p value <0.0001.

As previously discussed, CXCL13 promotes lymphoid neogenesis, resulting in the creation of ectopic lymphoid-like structures in non-lymphoid organs, referred to as TLSs^23–25^. These structures are frequently present in non-lymphoid tissues during chronic inflammatory states, such as infection, cancer, and autoimmune diseases^26^. The presence of TLSs in CRC correlates with infiltration of TFH cells and B cells, leading to protection against tumor recurrence^27^ and improved patient outcomes^20^. TLSs in this cohort were characterized by analyzing three gene signatures (see Material and Methods and Supplementary Table S6) Among treated patients, higher scores for the three gene signatures were correlated with improved OS. Furthermore, an association between TH1+B score and *CXCL13* expression was observed when comparing TH1+B scores between groups of treated patients with high and low CXCL13 gene expression (Wilcoxon Rank Sum Test, p-value 5e-4) (Figure 7D).

Finally, we considered the feasibility of reproducing our findings in a different cohort of patients. To achieve this, we employed serial serum samples which corresponded to the 36 patients enrolled in the control arm of the METIMMOX study and that were collected at 0, 4, 8, 12, and 16 weeks along with post-treatment samples (see materials and methods). CXCL13 concentrations in these samples were measured by ELISA, resulting in a median value of 77.09 pg/ml in baseline samples, akin to the result we previously obtained by Luminex. CXCL13 values progressively increased until the eighth week following treatment initiation. However, at 12 and 16 weeks, values then decreased, with the lowest value observed at 16 weeks (Supplementary Figure S4A). The increase in CXCL13 at the different time points in relation to the baseline sample value had a favorable impact on OS, being statistically significant only at the end of treatment (Supplementary Figure S4B and S4C). Similarly, the effect on PFS was positive but not statistically significant in any instance (Supplementary Figure S4B and S4D).

## 4. Discussion

CRC continues to be one of the primary causes for cancer-related fatalities across the globe. While improvements have been made recently in terms of molecular classification, diagnosis, and the introduction of advanced therapeutic regimens, there is still a requirement to develop new treatments and identify biomarkers that have the potential to enable tailored treatment approaches, consequently improving the prognosis and quality of life for patients. This study concentrates on investigating the CXC chemokine family as promising prognostic biomarkers in the serum of patients with metastatic CRC. Sequential peripheral blood samples were taken before treatment, at the time of response assessment, and at patient progression or last follow-up. Overall, increasing levels of CXCL13 was linked to response, better overall survival and progression-free survival in our cohort of patients. Increased CXCL13 expression in the primary tumors of these patients was significantly associated with an increased pattern of B and CD8+ T lymphocyte infiltration. Comparable findings were procured using *in silico* data from a similar cohort, where we were also able to portray the correlation between CXCL13, the presence of TLSs and the correlation of both with prognosis. To the best of our knowledge, this is the first study to report a correlation between serum CXCL13 and prognosis in patients with CRC who were treated with oxaliplatin.

Cancer is a systemic disease in which the inflammatory response is critical at every stage, including treatment response and progression^28^. In this context, analyzing the blood content of cancer patients can provide a non-invasive and time-specific tool that reflects the complete tumor burden. Such an approach offers a more comprehensive understanding of disease progression compared to methods that investigate only a part of a single lesion. Therefore, several studies have concentrated on the prospective function of chemokines as predictive and/or prognostic biomarkers in the blood of cancer patients^29^. These biomarkers are effortlessly detected in the blood of patients, are involved in inflammation and cancer, and can be impacted by antineoplastic therapy.

Our findings show that most protumoral chemokines followed a V-shaped trend, consisting of a decrease in mean concentration at EVAR compared to PRET and an increase at LFUP compared to EVAR. This shape was flatter in individuals who did not respond. In the case of CXCL8, it displayed a distinct pattern difference between those who responded and those who did not: in the former group, levels of CXCL8 decreased slightly in the EVAR sample and subsequently increased slightly in the LFUP sample, whereas in the latter group, a marked and progressive increase was detected in both the EVAR and LFUP samples. These findings are consistent with previous research conducted by our group, demonstrating that resistance to oxaliplatin in cell lines is contingent upon the activation of the NF-kB pathway. This subsequently results in heightened expression and secretion of CXCL8^10^. Furthermore, other studies have reported increased levels of CXCL8 in blood samples from NSCLC patients that do not respond to tyrosine kinase inhibitors or anti-PD1 therapy and in blood samples from melanoma patients that do not respond to anti-PD1 therapy^30,31^. These findings may be related to the fact that patients with liver and lung metastases had higher baseline levels of CXCL8 in comparison to those with metastases only in liver, lung or in other sites. In addition, patients with multiple metastases also had elevated levels of CXCL8, suggesting a strong relationship between this cytokine and tumor burden, consistent with previous research^32^.

The sole chemokine exhibiting an opposing behavior was CXCL13, as it rose in responders and progressively dropped in non-responders in the EVAR and LFUP samples. The increase of CXCL13 in EVAR sample was concurrently and statistically significantly linked with a decrease in risk of progression and death and was independent of surgery and other prognostic factors. On the other hand, patients in whom this chemokine decreased in the EVAR sample had a poorer prognosis. These findings suggest that effective treatment leads to heightened levels of CXCL13 in the peripheral blood, indicating a long-term immune response that is associated with improved PFS and OS^33^. This differs from the other chemokines, which would exhibit a more local, tumor-level effect. All patients enrolled in the study received treatment containing oxaliplatin, which is known to induce immunogenic cell death (ICD)^34^. The ICD process involves the secretion and exposure of specific membrane molecules, such as calreticulin, by tumor cells, which bring about DC maturation, leading to the activation of tumor-specific T lymphocytes that result in the obliteration of tumor cells. DCs have the ability to relocate to secondary or tertiary lymphoid organs, where they present tumor antigens to T cells, triggering an anti-tumor response^35,36^. CXCL13 is one of the chemokines that contribute to the development of an immune response in TLSs. It plays a vital role in attracting B cells and TFH, thereby promoting the generation of TLSs. Recent data indicate that TLSs have the potential to induce or reactivate anti-tumor immunity. In several types of solid organ tumors, including CRC, the presence of TLSs is often correlated with a more favorable prognosis^37^.

We hypothesized that the increase in CXCL13 seen in our patients could be due to the mobilization of this chemokine at a systemic level as a consequence of the treatment. Nonetheless, due to the inaccessibility of material from treated metastases, we were unable to confirm this hypothesis in our patients. However, we have shown that in primary tumors, higher expression of the *CXCL13* gene correlates with more immunogenic TME and a better prognosis. Moreover, using *in silico* data, we have shown that increased *CXCL13* expression in liver metastases from CRC patients treated with oxaliplatin in the neoadjuvant setting, is associated with an immunogenic milieu, the presence of TLS and improved OS.

Finally, we studied the dynamics of CXCL13 changes in blood samples from an independent cohort of patients. We observed a similar trend, but the results were not statistically significant. These results may be explained by limitations such as the small sample size or differences in the type of treatment received (Nordic FLOX scheme in this case).

In conclusion, we have demonstrated that oxaliplatin-based first-line treatment leads to changes in blood levels of chemokines CXCL1, CXCL2, CXCL5, CXCL6, CXCL8, CXCL9, CXCL10, CXCL12, CXCL13, and CXCL16 in CRC patients. Notably, an increase in CXCL13 compared with baseline after 12 cycles of treatment is associated with treatment response, improved OS and PFS, while a decrease is associated with non-response and worse prognosis. Our results suggest that the rise in blood CXCL13 may indicate a more immunogenic tumor microenvironment with a higher presence of TLSs. However, our study has limitations, including the loss of follow-up samples, the absence of metastasis samples for comparison, the lack of immunophenotyping in PBMCs, and the use of multidrug chemotherapy regimens. Therefore, further research is necessary to determine whether CXCL13 can serve as a reliable predictive biomarker in CRC.

## 5. Conclusions

The chemokines CXCL1, CXCL2, CXCL5, CXCL6, CXCL8, CXCL9, CXCL10, CXCL11, CXCL12, CXCL13, and CXCL16 may be detected in the blood of CRC patients undergoing first-line oxaliplatin treatment, as assessed by Luminex. Notably, these chemokine levels display dynamic changes depending on the timing of sample collection.

Specifically, the increase in blood levels of CXCL13 correlate with a more favorable prognosis among patients undergoing first-line oxaliplatin-based regimens. Our findings indirectly propose that these observed changes in blood levels may be indicative of a treatment-induced immune response. Moreover, they appear to be linked to a more immunogenic tumor microenvironment and the presence of TLSs, both of which are recognized as positive prognostic factors.

In light of these insights, it is conceivable that blood levels of CXCL13, in conjunction with the assessment of TLSs in the tumor, could function as a valuable prognostic biomarker for CRC patients undergoing first-line oxaliplatin-based treatment.

## Declarations section

## Data Availability

All data produced in the present study are available upon reasonable request to the authors

## Acknowledgements

We thank Dr Marco Antonio Fernández from the IGTP Flow Citometry Core Facility for his contribution to this publication

We want to particularly acknowledge the patients and the IGTP-HUGTP Biobank integrated in the Spanish National Biobanks Network of Instituto de Salud Carlos III (PT13/0010/0009) and Tumour Bank Network of Catalonia for its collaboration.

## Ethics approval and consent to participate

The study received approval from the Ethics Committee of Germans Trias I Pujol University Hospital under the internal reference number IP-16-115. All patients received proper information and signed the corresponding informed consent

## Consent for publication

Not applicable

## Data availability statement

The data and the datasets used and/or analyzed during the current study are available from the corresponding author on reasonable request.

## 6. References

1. Sung, H., et al. Global Cancer Statistics 2020: GLOBOCAN Estimates of Incidence and Mortality Worldwide for 36 Cancers in 185 Countries. CA Cancer J Clin 71, 209–249 (2021).

2. Cervantes, A. et al. Metastatic colorectal cancer: ESMO Clinical Practice Guideline for diagnosis, treatment and follow-up. Annals of Oncology 34, 10–32 (2023).

3. Koizumi, K., Hojo, S., Akashi, T., Yasumoto, K. & Saiki, I. Chemokine receptors in cancer metastasis and cancer cell-derived chemokines in host immune response. Cancer Sci 98, 1652–1658 (2007).

4. Vandercappellen, J., Van Damme, J. & Struyf, S. The role of CXC chemokines and their receptors in cancer. Cancer Lett 267, 226–244 (2008).

5. Rajagopal, S., Rajagopal, K. & Lefkowitz, R. J. Teaching old receptors new tricks: biasing seven-transmembrane receptors. Nat Rev Drug Discov 9, 373–386 (2010).

6. Nagarsheth, N., Wicha, M. S. & Zou, W. Chemokines in the cancer microenvironment and their relevance in cancer immunotherapy. Nat Rev Immunol 17, 559–572 (2017).

7. Bikfalvi, A. & Billottet, C. The CC and CXC chemokines: major regulators of tumor progression and the tumor microenvironment. American Journal of Physiology-Cell Physiology 318, C542–C554 (2020).

8. Acharyya, S. et al. A CXCL1 paracrine network links cancer chemoresistance and metastasis. Cell 150, 165–78 (2012).

9. Schimek, V. et al. Tumour cell apoptosis modulates the colorectal cancer immune microenvironment via interleukin-8-dependent neutrophil recruitment. Cell Death Dis 13, 113 (2022).

10. Ruiz de Porras, V., et al. Curcumin mediates oxaliplatin-acquired resistance reversion in colorectal cancer cell lines through modulation of CXC-Chemokine/NF-κB signalling pathway. Sci Rep 6, 24675 (2016).

11. Eide, P. W. et al. Metastatic heterogeneity of the consensus molecular subtypes of colorectal cancer. NPJ Genom Med 6, 59 (2021).

12. Moosavi, S. H. et al. De novo transcriptomic subtyping of colorectal cancer liver metastases in the context of tumor heterogeneity. Genome Med 13, 143 (2021).

13. Coppola, D. et al. Unique ectopic lymph node-like structures present in human primary colorectal carcinoma are identified by immune gene array profiling. Am J Pathol 179, 37–45 (2011).

14. Messina, J. L. et al. 12-Chemokine gene signature identifies lymph node-like structures in melanoma: potential for patient selection for immunotherapy? Sci Rep 2, 765 (2012).

15. Hennequin, A. et al. Tumor infiltration by Tbet+ effector T cells and CD20+ B cells is associated with survival in gastric cancer patients. Oncoimmunology 5, e1054598 (2016).

16. Barbie, D. A. et al. Systematic RNA interference reveals that oncogenic KRAS-driven cancers require TBK1. Nature 462, 108–12 (2009).

17. Becht, E. et al. Estimating the population abundance of tissue-infiltrating immune and stromal cell populations using gene expression. Genome Biol 17, 218 (2016).

18. Meltzer, S. et al. Early radiologic signal of responsiveness to immune checkpoint blockade in microsatellite-stable/mismatch repair-proficient metastatic colorectal cancer. Br J Cancer 127, 2227–2233 (2022).

19. Bindea, G. et al. Spatiotemporal Dynamics of Intratumoral Immune Cells Reveal the Immune Landscape in Human Cancer. Immunity 39, 782–795 (2013).

20. Sautès-Fridman, C., Petitprez, F., Calderaro, J. & Fridman, W. H. Tertiary lymphoid structures in the era of cancer immunotherapy. Nat Rev Cancer 19, 307–325 (2019).

21. Newman, A. M. et al. Determining cell type abundance and expression from bulk tissues with digital cytometry. Nat Biotechnol 37, 773–782 (2019).

22. CIBERSORTx. https://cibersortx.stanford.edu/.

23. Luther, S. A., Lopez, T., Bai, W., Hanahan, D. & Cyster, J. G. BLC Expression in Pancreatic Islets Causes B Cell Recruitment and Lymphotoxin-Dependent Lymphoid Neogenesis. Immunity 12, 471–481 (2000).

24. Dieu-Nosjean, M.-C., Goc, J., Giraldo, N. A., Sautès-Fridman, C. & Fridman, W. H. Tertiary lymphoid structures in cancer and beyond. Trends Immunol 35, 571–580 (2014).

25. Nerviani, A. & Pitzalis, C. Role of chemokines in ectopic lymphoid structures formation in autoimmunity and cancer. J Leukoc Biol 104, 333–341 (2018).

26. Schumacher, T. N. & Thommen, D. S. Tertiary lymphoid structures in cancer. Science (1979) 375, (2022).

27. Bindea, G., Mlecnik, B., Angell, H. K. & Galon, J. The immune landscape of human tumors. Oncoimmunology 3, e27456 (2014).

28. Grivennikov, S. I., Greten, F. R. & Karin, M. Immunity, Inflammation, and Cancer. Cell 140, 883–899 (2010).

29. Heras, S. C. las & Martínez-Balibrea, E. CXC family of chemokines as prognostic or predictive biomarkers and possible drug targets in colorectal cancer. World J Gastroenterol 24, 4738–4749 (2018).

30. Sanmamed, M. F. et al. Changes in serum interleukin-8 (IL-8) levels reflect and predict response to anti-PD-1 treatment in melanoma and non-small-cell lung cancer patients. Ann Oncol 28, 1988–1995 (2017).

31. Angeles, A. K. et al. Integrated circulating tumour DNA and cytokine analysis for therapy monitoring of ALK-rearranged lung adenocarcinoma. Br J Cancer 129, 112–121 (2023).

32. Alfaro, C. et al. Interleukin-8 in cancer pathogenesis, treatment and follow-up. Cancer Treat Rev 60, 24–31 (2017).

33. Galluzzi, L., Buqué, A., Kepp, O., Zitvogel, L. & Kroemer, G. Immunological Effects of Conventional Chemotherapy and Targeted Anticancer Agents. Cancer Cell 28, 690–714 (2015).

34. Galluzzi, L. et al. Molecular mechanisms of cell death: recommendations of the Nomenclature Committee on Cell Death 2018. Cell Death Differ 25, 486– 541 (2018).

35. Wang, Y. et al. The consensus on the monitoring, treatment, and prevention of leukemia relapse after allogeneic hematopoietic stem cell transplantation in China. Cancer Lett 438, 63–75 (2018).

36. Pol, J. G., Le Naour, J. & Kroemer, G. FLT3LG - a biomarker reflecting clinical responses to the immunogenic cell death inducer oxaliplatin. Oncoimmunology 9, (2020).

37. Fridman, W. H. et al. Tertiary lymphoid structures and B cells: An intratumoral immunity cycle. Immunity 56, 2254–2269 (2023).

