## Supplementary materials for "Changes in serum CXCL13 levels are associated with outcomes of Colorectal Cancer Patients Undergoing First-Line Oxaliplatin-Based Treatment"

**Cabrero-de las Heras, S et al. Supplementary materials.**

1. **Supplementary Figures' legends**

**Supplementary Figure S1**. **Association of all chemokines with different clinical features** Box plots presenting the median concentration (horizontal line) and standard deviation (SD, vertical line) of the chemokines under study in each study sample, categorized by number of metastasis (A), performance status (B), sex (C) and primary tumor site (D). The p-values resulting from Kruskal-Wallis test are provided at the top of each plot.

**Supplementary Figure S2. Dynamics of CXCL13 in the LFUP samples according to increase or decrease in the EVAR sample.** The box plots show the median CXCL13 values (horizontal line) and the SD (vertical line) in each of the samples obtained. Patients were divided into two groups, according to the increase or decrease of CXCL13 in the EVAR sample with respect to PRET. A) Results corresponding to all cases. B) Results after excluding patients who had not progressed at the time of LFUP sample extraction. The p values corresponding to the Wilcoxon signed-rank test are shown at the top of each panel.

**Supplementary Figure S3. Correlation of CXCL13 expression and genes associated with immune cell populations in tumors.** Scatter plots illustrating the correlation between *CXCL13* gene expression and representative genes of regulatory T cells (Tregs), Natural Killer cells (NKs), Dendritic cells (DCs), Cytotoxic T Lymphocytes (Cytotoxic) and Follicular T helper cells (TFH), as indicated. The legend provides p and r values corresponding to the Spearman test.

**Supplementary Figure S4**. **Results of the METIMMOX cohort.** A) Line plot showing mean CXCL13 serum levels at baseline, 4, 8, 12, 16 weeks and at the end of treatment. B) OS and PFS according to increase or decrease of CXCL13 levels at indicated time points. Hazard Ratios (HR) correspond to category "increase", being "decrease" the reference category; p values, HR, and 95% Confidence Intervals (CI) correspond to univariate[1] and multivariate[2] COX regression models (adjusted by age, sex, and performance status). Statistically significant results ($p < 0.05$) are highlighted in bold. C) Kaplan-Meier plots showing PFS and OS of patients split according to an increase or decrease of CXCL13 at the end of treatment.

#### 2. Supplementary Figures

##### Supplementary Figure S1

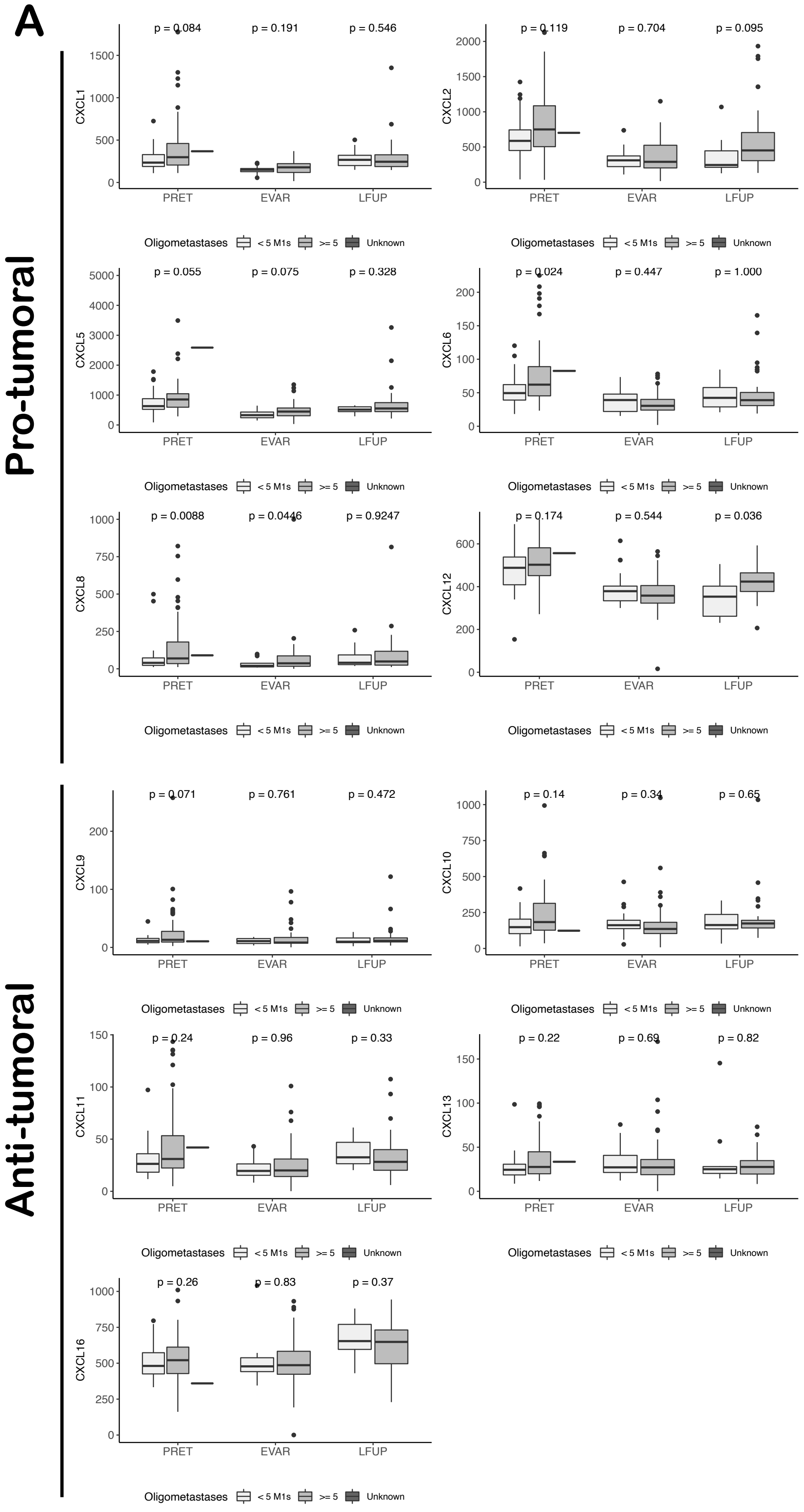

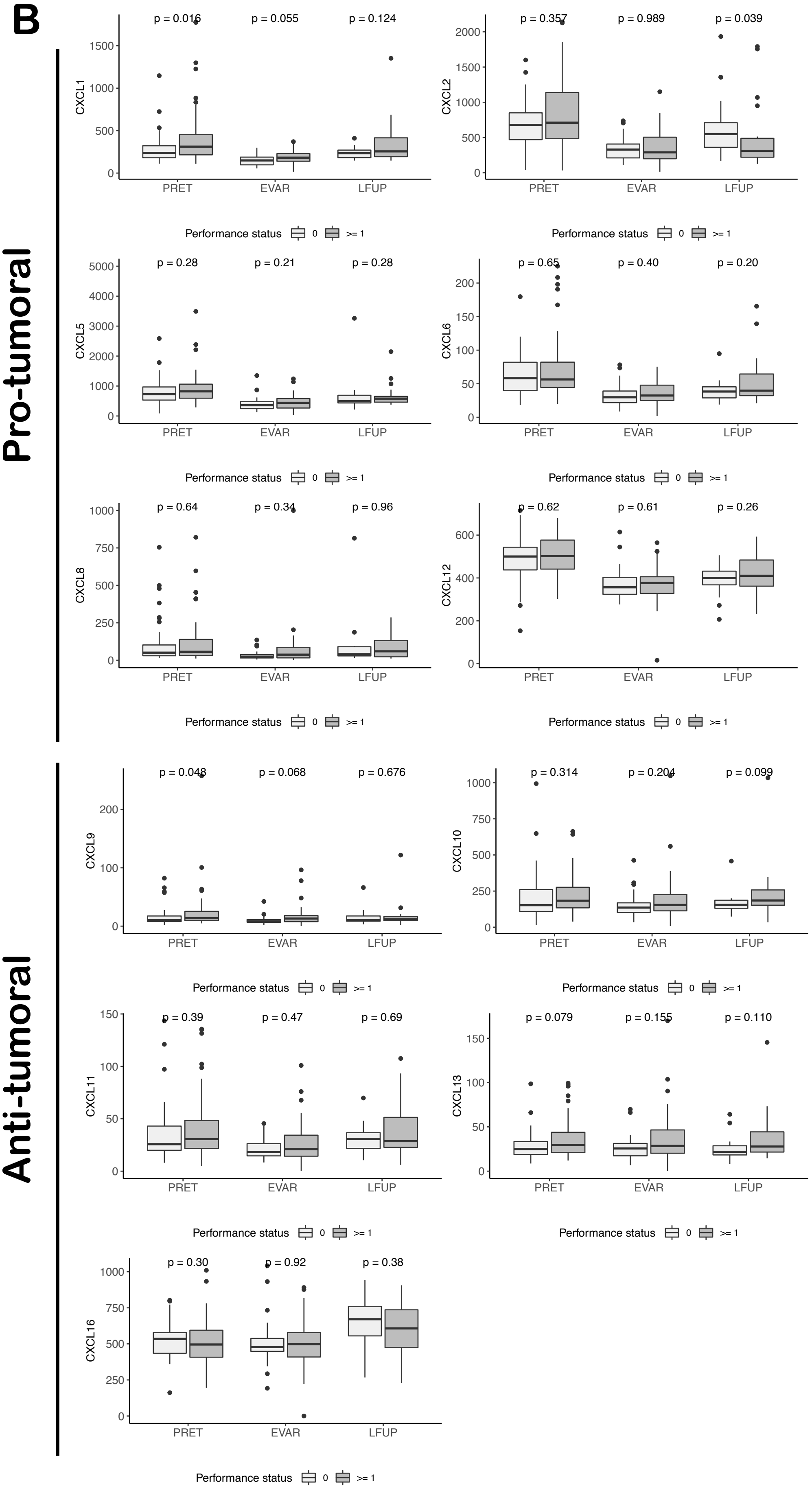
B
Pro-tumoral
Anti-tumoral
CXCL1
p = 0.016
p = 0.055
p = 0.124
CXCL2
p = 0.357
p = 0.989
p = 0.039
CXCL5
p = 0.28
p = 0.21
p = 0.28
CXCL6
p = 0.65
p = 0.40
p = 0.20
CXCL8
p = 0.64
p = 0.34
p = 0.96
CXCL12
p = 0.62
p = 0.61
p = 0.26
CXCL9
p = 0.048
p = 0.068
p = 0.676
CXCL10
p = 0.314
p = 0.204
p = 0.099
CXCL11
p = 0.39
p = 0.47
p = 0.69
CXCL13
p = 0.079
p = 0.155
p = 0.110
CXCL16
p = 0.30
p = 0.92
p = 0.38
PRET
EVAR
LFUP
Performance status
0
>= 1

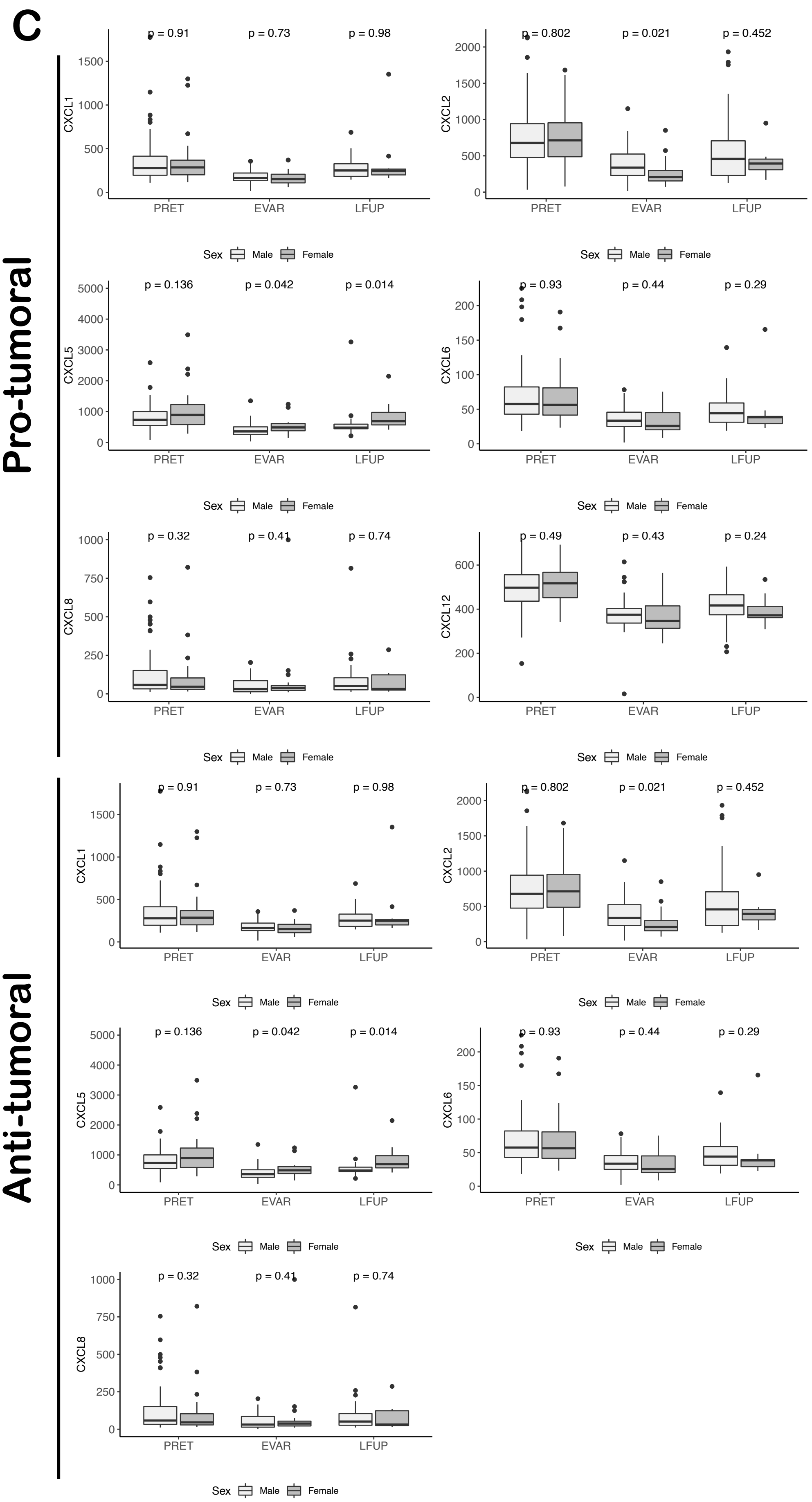
C
Pro-tumoral
Anti-tumoral
CXCL1
CXCL2
CXCL5
CXCL6
CXCL8
CXCL12
PRET
EVAR
LFUP
Sex
Male
Female
p = 0.91
p = 0.73
p = 0.98
p = 0.802
p = 0.021
p = 0.452
p = 0.136
p = 0.042
p = 0.014
p = 0.93
p = 0.44
p = 0.29
p = 0.32
p = 0.41
p = 0.74
p = 0.49
p = 0.43
p = 0.24

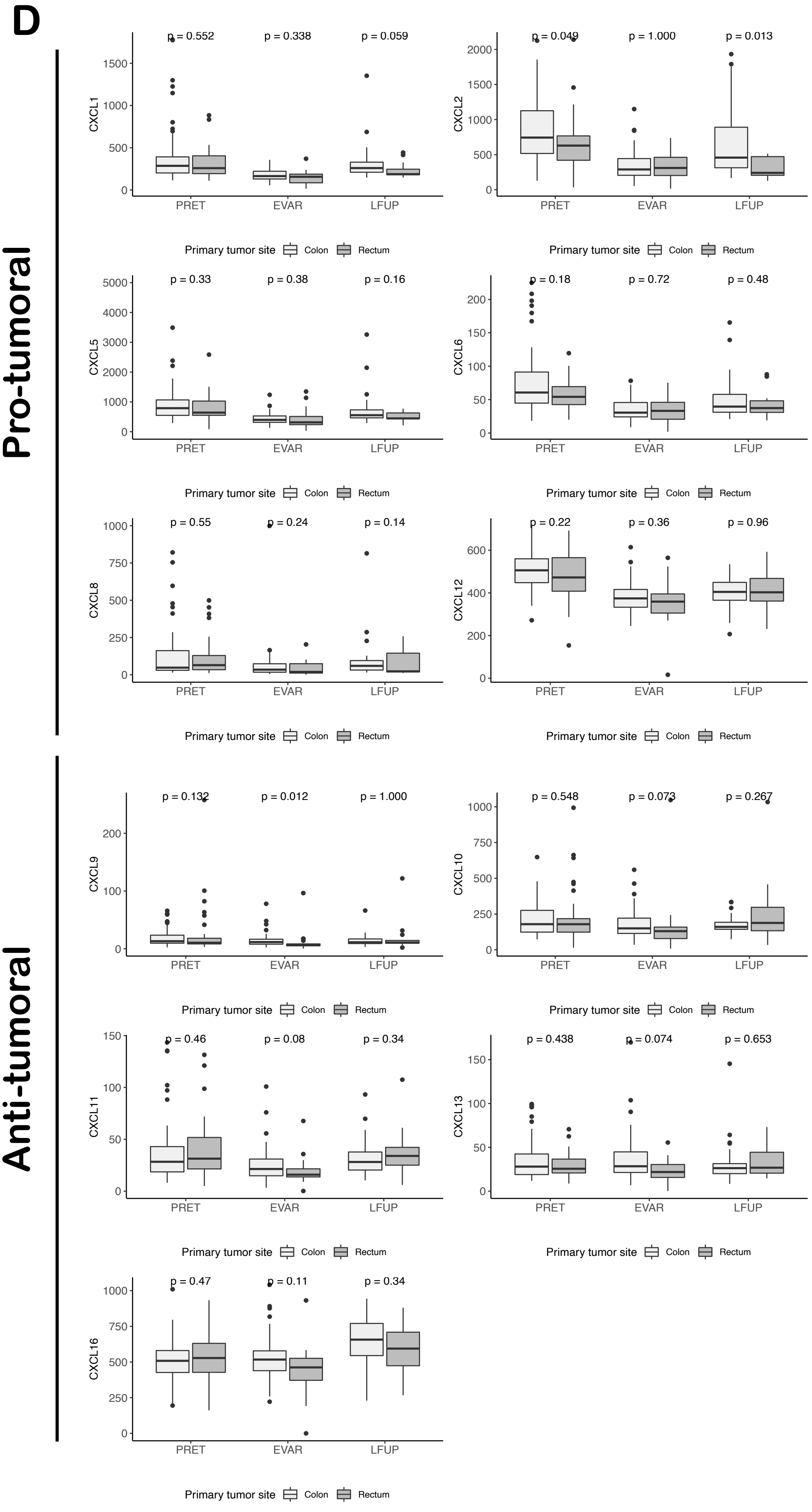
D
Pro-tumoral
Anti-tumoral
CXCL1
p = 0.552
p = 0.338
p = 0.059
CXCL2
p = 0.049
p = 1.000
p = 0.013
CXCL5
p = 0.33
p = 0.38
p = 0.16
CXCL6
p = 0.18
p = 0.72
p = 0.48
CXCL8
p = 0.55
p = 0.24
p = 0.14
CXCL12
p = 0.22
p = 0.36
p = 0.96
CXCL9
p = 0.132
p = 0.012
p = 1.000
CXCL10
p = 0.548
p = 0.073
p = 0.267
CXCL11
p = 0.46
p = 0.08
p = 0.34
CXCL13
p = 0.438
p = 0.074
p = 0.653
CXCL16
p = 0.47
p = 0.11
p = 0.34
PRET
EVAR
LFUP
Primary tumor site
Colon
Rectum

#### Supplementary Figure S2

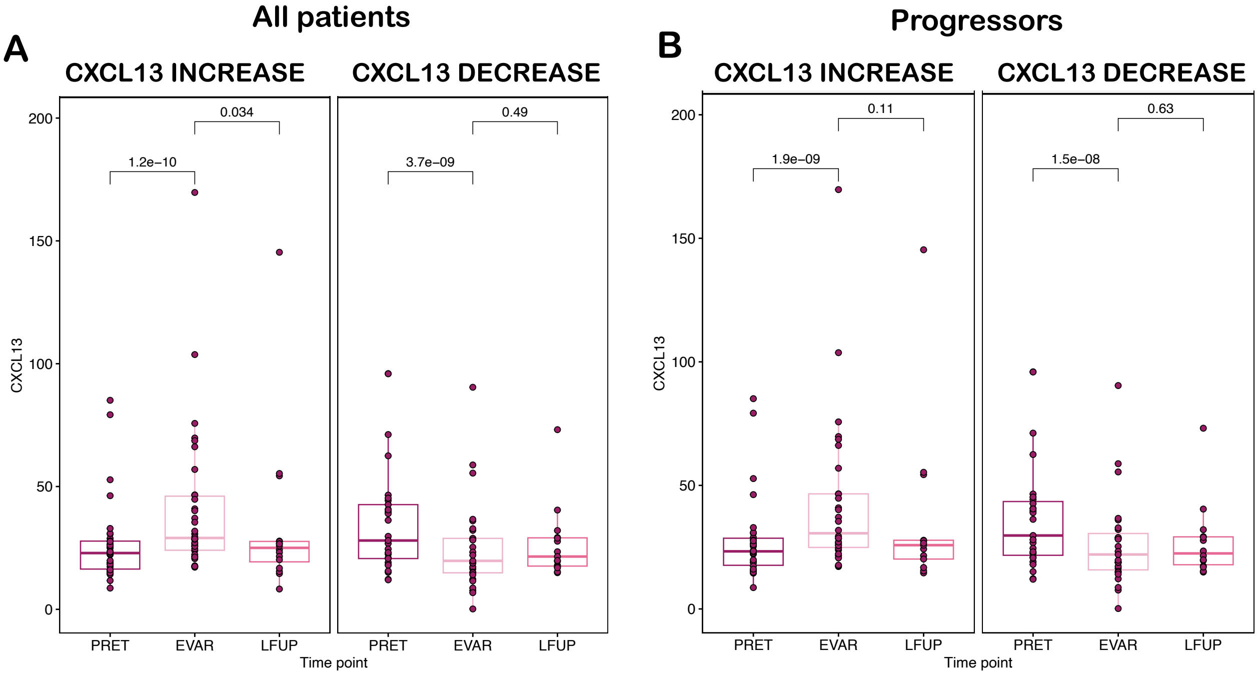

#### Supplementary Figure S3

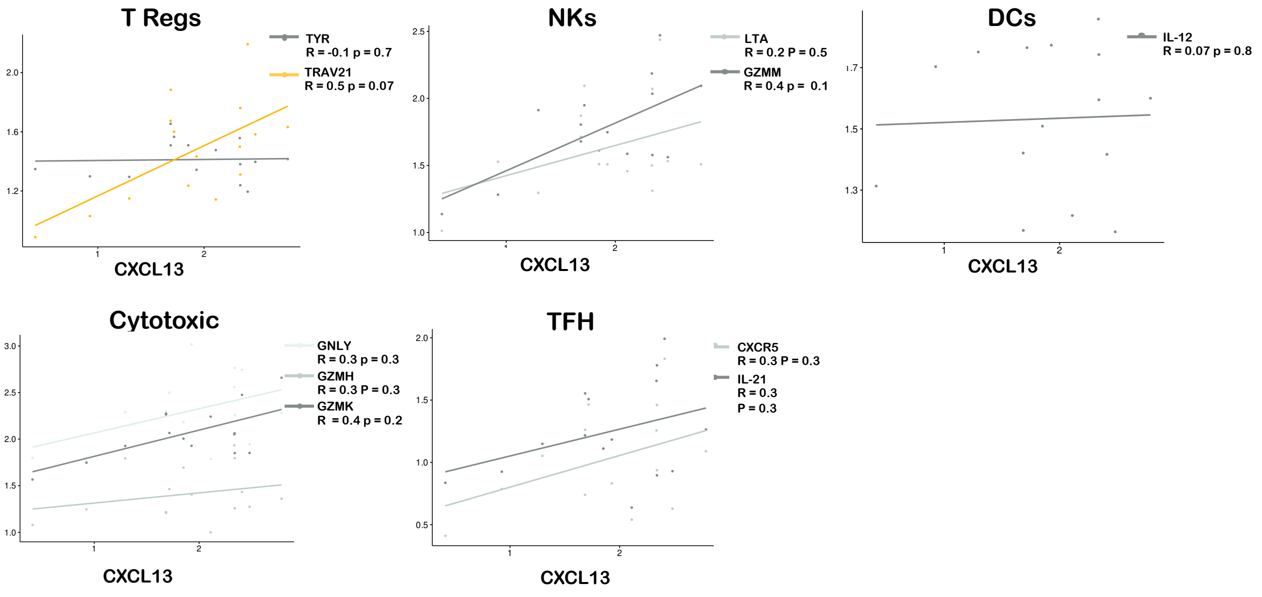

### Supplementary Figure S4

A

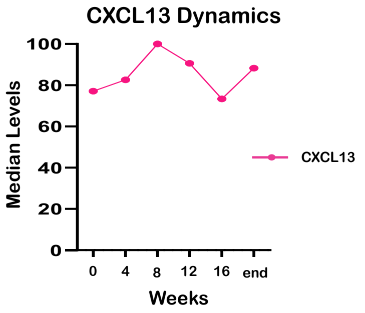


B

| CXCL13 Dynamic changes | | | | | | |
|---|---|---|---|---|---|---|
| | | Overall Survival | | Progression-Free Survival | | |
| Change at | Change | HR (95% CI) p-value[1] | HR (95% CI) p-value[2] | HR (95% CI) p-value[1] | HR (95% CI) p-value[2] | N |
| 4 weeks | Decrease / Increase | 0.91 (0.32 - 2.62) 0.86 | 0.64 (0.19 - 2.21) 0.48 | 0.69 (0.2 - 2.41) 0.69 | 0.53 (0.12 - 2.25) 0.39 | 33 |
| 8 weeks | Decrease / Increase | 0.96 (0.47 - 1.97) 0.92 | 0.75 (0.28 - 1.99) 0.56 | 0.88 (0.45 - 1.73) 0.72 | 0.85 (0.39 - 1.87) 0.69 | 28 |
| 12 weeks | Decrease / Increase | 0.83 (0.38 - 1.82) 0.64 | 0.49 (0.14 - 1.68) 0.26 | 0.84 (0.45 - 1.55) 0.57 | 0.69 (0.34 - 1.42) 0.32 | 23 |
| 16 weeks | Decrease / Increase | 1.60 (0.44 - 5.84) 0.48 | 1.33 (0.32 - 5.48) 0.69 | 3.88 (0.71 - 21.16) 0.12 | 2.64 (0.43 - 16.25) 0.30 | 17 |
| End of treatment | Decrease / Increase | 0.55 (0.26 - 1.16) 0.12 | **0.29 (0.11 - 0.76) 0.01** | 0.73 (0.36 - 1.46) 0.38 | 0.48 (0.19 - 1.17) 0.11 | 33 |

C

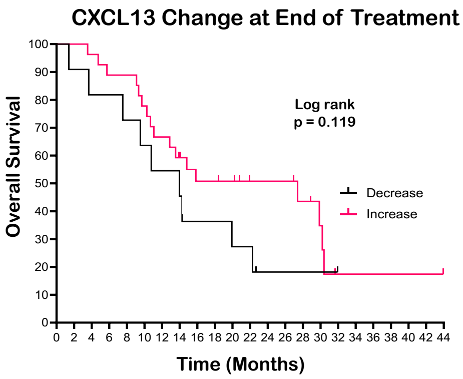


D

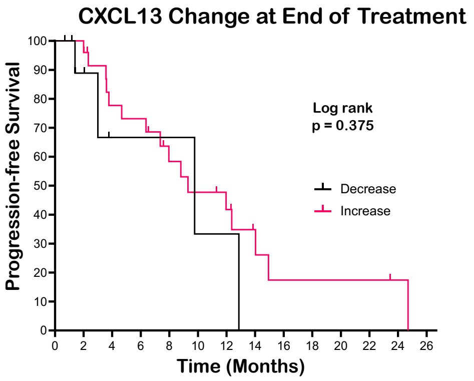

### 3. Supplementary Tables

**Supplementary Table S1. First-Line regimens**

| CHEMOTHERAPY | SYSTEMIC TREATMENT REGIMEN | FREQUENCY |
|---|---|---|
| **Oxaliplatin (OXA)** | | |
| FOLFOX | LV: 400mg/m² on day 1 + | Every 2 weeks |
| | 5FU: 400mg/m² bolus on day 1, 5FU 2400mg/m² infusion on 46h + | |
| | OXA: 85mg/m² on day 1 | |
| CAPOX | Cape: 1000mg/m² every 12h oral day 1 to day 14 + | Every 3 weeks |
| | OXA: 130mg/m² day 1 | |
| **Bevacizumab** | | |
| FOLFOX-Bevacizumab | Combined with FOLFOX regimen | Every 2 weeks |
| | Beva: 5 mg/kg | |
| CAPOX-Bevacizumab | Combined with CAPOX regimen | Every 3 weeks |
| | Beva: 7,5 mg/kg | |
| **Panitumumab** | | |
| FOLFOX-Panitumumab | Combined with FOLFOX regimen + | Every 2 weeks |
| | Pani: 6 mg/kg | |
| **Cetuximab** | | |
| FOLFOX-Cetuximab | Combined with FOLFOX regimen + | Weekly |
| | Cetux: 400 mg/m² first infusion, followed by 250 mg/m² or | or |
| | 500 mg/m² over 2h on day 1 (preferred) | Every 2 weeks (preferred) |
| CAPOX-Cetuximab | Combined with CAPOX regimen + | Weekly |
| | Cetux: 400 mg/m² first infusion, followed by 250 mg/m² | |

**Supplementary Table 2. Probes for Nanostring Analysis**

| Probe Name | NS Probe ID | Class Name | Analyte Type | Species Name | Target Sequence |
|---|---|---|---|---|---|
| CD19 | XM_011545981.1:713 | Endogenous | mRNA | Homo sapiens | CACCCCAAGGGGCCTAAGTCATTGCTGAGCCTAGAGCTGAAGGACGATCGCCCGGCCAGAGATATGTGGGTAATGGAGACGGGTCTGTTGTTGCCCCGGG |
| CD40 | NM_001250.4:196 | Endogenous | mRNA | Homo sapiens | GTGCCAGCCAGGACAGAAACTGGTGAGTGACTGCACAGAGTTCACTGAAACGGAATGCCTTCCTTGCGGTGAAAGCGAATTCCTAGACACCTGGAACAGA |
| CD79A | NM_001783.3:695 | Endogenous | mRNA | Homo sapiens | AACGAGAAGCTCGGGTTGGATGCCGGGGATGAATATGAAGATGAAAACCTTTATGAAGGCCTGAACCTGGACGACTGCTCCATGTATGAGGACATCTCCC |
| CD8A | NM_001768.5:1320 | Endogenous | mRNA | Homo sapiens | GCTCAGGGCTCTTTCCTCCACACCATTCAGGTCTTTCTTTCCGAGGCCCCTGTCTCAGGGTGAGGTGCTTGAGTCTCCAACGGCAAGGGAACAAGTACTT |
| CD8B | NM_172099.2:439 | Endogenous | mRNA | Homo sapiens | TCAGCTGAGTGTGGTTGATTTCCTTCCCACCACTGCCCAGCCCACCAAGAAGTCCACCCTCAAGAAGAGAGTGTGCCGGTTACCCAGGCCAGAGACCCAG |
| CRTAM | NM_019604.2:775 | Endogenous | mRNA | Homo sapiens | AAGAGAAAGAACAAACCACTCAAGATCCTGACTTGACCACCGAAGCAAATCCTCAGTATTTAGGACTGGCAAGAAAGAAAAGTGGCATCCTGCTGCTCAC |
| CXCL13 | NM_006419.2:210 | Endogenous | mRNA | Homo sapiens | AGACGCTTCATTGATCGAATTCAAATCTTGCCCCGTGGGAATGGTTGTCCAAGAAAAGAAATCATAGTCTGGAAGAAGAACAAGTCAATTGTGTGTGTGG |
| CXCR5 | NM_001716.3:2618 | Endogenous | mRNA | Homo sapiens | ACGTCCCTTTTTTCTCTGAGTATCTCCTCGCAAGCTGGGTAATCGATGGGGGAGTCTGAAGCAGATGCAAAGAGGCAAGAGGCTGGATTTTGAATTTTCT |
| GNLY | NM_012483.2:880 | Endogenous | mRNA | Homo sapiens | TGCCGGCTCCTCGCTTCCTCGATCCAGAATCCACTCTCCAGTCTCCCTCCCCTGACTCCCTCTGCTGTCCTCCCCTCTCACGAGAATAAAGTGTCAAGCA |
| GZMB | NM_004131.4:733 | Endogenous | mRNA | Homo sapiens | ATGGCATGCCTCCACGAGCCTGCACCAAAGTCTCAAGCTTTGTACACTGGATAAAGAAAACCATGAAACGCTACTAACTACAGGAAGCAAACTAAGCCCC |
| GZMH | NM_033423.3:705 | Endogenous | mRNA | Homo sapiens | AAAAAAGGGACACCTCCAGGAGTCTACATCAAGGTCTCACACTTCCTGCCCTGGATAAAGAGAACAATGAAGCGCCTCTAACAGCAGGCATGAGACTAAC |
| GZMK | NM_002104.2:700 | Endogenous | mRNA | Homo sapiens | CTGTAAAGGTGTCTTCCACGCTATAGTCTCTGGAGGTCATGAATGTGGTGTTGCCACAAAGCCTGGAATCTACACCCTGTTAACCAAGAAATACCAGACT |
| GZMM | NM_005317.3:704 | Endogenous | mRNA | Homo sapiens | CTGTCCTTCAGCTCCAGGGTCTGCACTGACATCTTCAAGCCTCCCGTGGCCACCGCTGTGGCGCCTTACGTGTCCTGGATCAGGAAGGTCACCGGCCGAT |
| HLA-DOB | NM_002120.3:230 | Endogenous | mRNA | Homo sapiens | ACGGGACAGAAAAGGTGCAGTTTGTGGTCAGATTCATCTTTAACTTGGAGGAGTATGTACGTTTCGACAGTGATGTGGGGATGTTTGTGGCATTGACCAA |
| HLA-DQA1 | NM_002122.3:261 | Endogenous | mRNA | Homo sapiens | GGTGGCCTGAGTTCAGCAAATTTGGAGGTTTTGACCCGCAGGGTGCACTGAGAAACATGGCTGTGGCAAAACACAACTTGAACATCATGATTAAACGCTA |
| IL-12 | NM_000882.2:775 | Endogenous | mRNA | Homo sapiens | CTTTCTAGATCAAAACATGCTGGCAGTTATTGATGAGCTGATGCAGGCCCTGAATTTCAACAGTGAGACTGTGCCACAAAAATCCTCCCTTGAAGAACCG |
| IL-21 | NM_021803.2:65 | Endogenous | mRNA | Homo sapiens | CATGGAGAGGATTGTCATCTGTCTGATGGTCATCTTCTTGGGGACACTGGTCCACAAATCAAGCTCCCAAGGTCAAGATCGCCACATGATTAGAATGCGT |
| LTA | NM_001159740.1:880 | Endogenous | mRNA | Homo sapiens | CCCATTCTGCCTCCATTCTGACCATTTCAGGGGTCGTCACCACCTCTCCTTTGGCCATTCCAACAGCTCAAGTCTTCCCTGATCAAGTCACCGGAGCTTT |
| MS4A1 | NM_152866.2:1412 | Endogenous | mRNA | Homo sapiens | CACATCTCTATCTGGCCTTTGCATGGAGTGACCATAGCTCCTTCTCTCTTACATTGAATGTAGAGAATGTAGCCATTGTAGCAGCTTGTGTTGTCACGCT |
| TRAV21 | ENST00000390449.1:149 | Endogenous | mRNA | Homo sapiens | CTATTTACAACCTCCAGTGGTTTAGGCAGGACCCTGGGAAAGGTCTCACATCTCTGTTGCTTATTCAGTCAAGTCAGAGAGAGCAAACAAGTGGAAGACT |
| TYR | NM_000372.4:1195 | Endogenous | mRNA | Homo sapiens | GGAACAATGTCCCAGGTACAGGGATCTGCCAACGATCCTATCTTCCTTCTTCACCATGCATTTGTTGACAGTATTTTTGAGCAGTGGCTCCGAAGGCACC |
| ACTB | NM_001101.2:1010 | Housekeeping | mRNA | Homo sapiens | TGCAGAAGGAGATCACTGCCCTGGCACCCAGCACAATGAAGATCAAGATCATTGCTCCTCCTGAGCGCAAGTACTCCGTGTGGATCGGCGGCTCCATCCT |
| HPRT1 | NM_000194.1:240 | Housekeeping | mRNA | Homo sapiens | TGTGATGAAGGAGATGGGAGGCCATCACATTGTAGCCCTCTGTGTGCTCAAGGGGGGCTATAAATTCTTTGCTGACCTGCTGGATTACATCAAAGCACTG |

**Supplementary Table 3. Patients' characteristics of the**

| VARIABLES | N = 36 |
|---|---|
| | N (%) |
| Age at Diagnosis (Med [IQR]) | 65 [19] |
| Sex | |
| Male | 22 (61%) |
| Female | 14 (39%) |
| Performance Status | |
| 0 | 19 (53%) |
| 1 | 17 (47%) |
| 2 | 0 |
| Primary Tumor Site | |
| Colon | 9 (25%) |
| Rectum | 9 (25%) |
| Unknown | 18 (50%) |
| Metastatic Site | |
| Liver | 7 (19%) |
| Lung | 0 |
| Liver+Lung | 13 (36%) |
| Other | 16 (45%) |
| Number of mestastases | |
| <5 Metastasis | 35 (97%) |
| ≥5 Metastasis | 1 (3%) |
| First-Line Treatment | |
| FLOX | 36 (100%) |
| CT combined with targeted therapy | 0 |
| Second Line | |
| Yes | 30 (83%) |
| No | 6 (17%) |
| Response Evaluation (Best) | |
| Complete response (CR) | 1 (3%) |
| Partial response (PR) | 20 (56%) |
| Stable disease (SD) | 8 (22%) |
| Progressive disease (PD) | 3 (8%) |
| Radical Surgery | |
| Yes | 5 (14%) |
| No | 31 (86%) |
| Resected Metastatic Organs | |
| Liver | 5 (14%) |
| Lung | 0 |
| Peritoneum | 0 |
| Microsatellite instability | |
| MSI | 0 |
| MSS | 36 (100%) |
| Unknown | 0 |
| BRAF and RAS mutation | |
| BRAF mut | 8 (22%) |
| NRAS mut | 4 (11%) |
| KRAS mut | 15 (42%) |
| WT | 9 (25%) |
| Immune System Known Alterations | |
| Autoimmune disease | |
| Patient with a translpant | |
| Major surgery last month | |
| Unknown | 36 (100%) |
| Exitus | |
| Yes | 34 (94%) |
| No | 2 (6%) |

**Supplementary Table S4. Clinical features and association with overall and progression-free survival**

| VARIABLES N (%) | ALL | OVERALL SURVIVAL (OS) | | PROGRESSION FREE SURVIVAL (PFS) | |
|---|---|---|---|---|---|
| | N = 104 | MEDIAN IN MONTHS (95% CI) | HR (95% CI) P VALUE | MEDIAN IN MONTHS (95% CI) | HR (95% CI) P VALUE |
| Sex | | | | | |
| Male | 74 (71.2%) | 29.4 (19.7-NR) | 1 | 11.5 (9.3-13.5) | 1 |
| Female | 30 (28.8%) | 20.3 (14.1-39.7) | 1.24 (0.72-2.13) 0.308 | 11 (8.7-15.7) | 1.27 (0.77-2.07) 0.348 |
| Performance Status | | | | | |
| 0 | 41 (39.4%) | 36 (30.1-NR) | 1 | 17.2 (12-24.5) | 1 |
| ≥1 | 63 (60.6%) | 17.8 (14.5-25.3) | 2.66 (1.47-4.81) 0.0005 | 10.1 (8.3-11.7) | 2.23 (1.33-3.73) 0.0008 |
| Primary Tumor Site | | | | | |
| Colon | 66 (63.5%) | 22.4 (15.1-30.6) | 1 | 10.6 (9.1-12.8) | 1 |
| Rectum | 38 (36.5%) | 36 (20.1-NR) | 0.58 (0.34-1.01, p=0.054) | 12.2 (10.1-18.7) | 0.84 (0.52-1.38) 0.498 |
| Metastastic Site | | | | | |
| Liver | 51 (49.0%) | 23 (18.4-NR) | 1 | 10.2 (8.3-14.5) | 1 |
| Lung | 11 (10.6%) | 36 (36-NR) | 0.49 (0.17-1.41) 0.187) | 14.1 (12.8-NR) | 0.73 (0.34-1.55) 0.408 |
| Liver + Lung | 31 (29.8%) | 23.1 (13.3-33.4) | 1.45 (0.82-2.56) 0.199) | 10.9 (8.7-13.6) | 1.20 (0.74-1.95) 0.451 |
| Other | 11 (10.6%) | 20.3 (12.6-NR) | 1.09 (0.45-2.61) 0.853) | 9.1 (6.5-NR) | 0.93 (0.45-1.91) 0.836 |
| Number of Metastasis | | | | | |
| < 5 Metastasis | 32 (30.8%) | NR (19.7-NR) | 1 | 12.8 (9.8-16.6) | 1 |
| ≥ 5 Metastasis | 72 (69.2%) | 20.3 (15.3-30.6) | 2.28 (1.23-4.24) 0.007 | 10.7 (8.7-12.7) | 1.07 (0.63-1.82) 0.801 |
| First Line Treatment | | | | | |
| Chemotherapy (CT) | 37 (35.6%) | 39.7 (17.1-NR) | 1 | 9.1 (7.7-23.2) | 1 |
| CT + Bevacizumab | 40 (38.5%) | 23.1 (19.7-33.4) | 1.49 (0.82-2.70) 0.188 | 12 (9.8-16.5) | 0.82 (0.48-1.43) 0.492 |
| CT + anti-EGFR | 27 (26.0%) | 30.6 (15.3-NR) | 0.98 (0.48-2.00) 0.950 | 11.7 (10.2-15.1) | 0.90 (0.48-1.69) 0.743 |
| Response Evaluation | | | | | |
| Complete or Partial Response | 64 (64%) | 33.4 (25.3-NR) | 1 | 12.7 (11-16.6) | 1 |
| Stable or Progressive disease | 36 (36%) | 13.8 (10.5-22.4) | 2.50 (1.5-4.18) 0.0001 | 7 (6.6-11.5) | 3.29 (2.00-5.41) 0.0001 |
| Radical Surgery | | | | | |
| Yes | 23 (22.1%) | NR | 1 | NR | NA |
| No | 81 (77.9%) | 19.6 (15.5-27.4) | 2.95 (1.65-5.28) <0.001 | 10.6 (9.1-12.4) | NA |
| Microsatellite instability | | | | | |
| MSI | 5 (4.8%) | 17.8 (14.1-NR) | 1 | 9.1 (6.6-NR) | 1 |
| MSS | 75 (72.1%) | 29.4 (22.4-39.7) | 0.69 (0.25-1.92) 0.473 | 11.7 (9.8-14.1) | 0.39 (0.15-1.01, p=0.052 |
| Unknown | 24 (23.1%) | - | - | - | - |
| BRAF and KRAS mutation | | | | | |
| BRAF mut | 4 (3.8%) | 16.2 (11.6-NR) | 1 | 9.1 (6.8-NR) | 1 |
| NRAS mut | 9 (8.7%) | 17.2 (7.5-NR) | 1.53 (0.39-5.98) 0.541 | 9.1 (7.5-NR) | 0.89 (0.23-3.4) 0.865 |
| KRAS mut | 46 (44.2%) | 25.3 (17.8-36) | 0.84 (0.26-2.79) 0.782 | 11.6 (8.9-14.8) | 0.43 (0.13-1.43) 0.168 |
| WT | 43 (41.3%) | 39.7 (20.1-NR) | 0.52 (0.15-1.74) 0.288 | 12.8 (10.6-15.7) | 0.43 (0.13-1.45) 0.171 |
| Unknown | 2 (1.9%) | - | - | - | - |

Median OS and PFS were estimated using the Kaplan-Meier method. Hazard Ratios (HR), 95% Confidence Intervals (CI) and p-values were obtained from the log-rank test.

NR = Not Reached; NA = Not Applicable as Not applicable time was censored at the date of surgery

**Supplementary Table S5. Overall and Progression-Free survival analysis according to CXC Chemokines' dynamics.**

| | **Overall Survival (OS)** | | | | | **Progression-Free Survival (PFS)** | | | | |
|---|---|---|---|---|---|---|---|---|---|---|
| | | **Unadjusted analysis** | | **IPW-adjusted analysis** | | | **Unadjusted analysis** | | **IPW-adjusted analysis** | |
| | **Median (95% CI)** | **HR (95% CI)** | **p-value** | **HR (95% CI)** | **p-value** | **Median (95% CI)** | **HR (95% CI)** | **p-value** | **HR (95% CI)** | **p-value** |
| **CXCL1** | | | | | | | | | | |
| Decrease | 27 (17 - NR) | 1 | | 1 | | 11 (9.9 - 15) | | | | |
| Increase | 27 (13 - NR) | 1.07 (0.42 - 2.77) | 0.88 | 1.05 (0.27 - 4.14) | 0.94 | 7.5 (6.5 - NR) | 1.68 (0.65 - 4.33) | 0.28 | 0.90 (0.15 - 5.54) | 0.91 |
| **CXCL2** | | | | | | | | | | |
| Decrease | 27 (16 - NR) | 1 | | 1 | | 11 (9.9 - 15) | | | | |
| Increase | 30 (18 - NR) | 0.89 (0.37 - 2.14) | 0.79 | 0.62 (0.20 - 1.94) | 0.42 | 8.1 (7.0 - NR) | 1.21 (0.50 - 2.88) | 0.67 | 0.93 (0.24 - 3.58) | 0.91 |
| **CXCL5** | | | | | | | | | | |
| Decrease | 25 (17 - NR) | 1 | | 1 | | 11 (9.8 - 14) | | | | |
| Increase | 30 (NR - NR) | 0.70 (0.17 - 2.94) | 0.63 | 0.85 (0.11 - 6.25) | 0.87 | NR (7.0 - NR) | 0.55 (0.13 - 2.28) | 0.41 | 0.61 (0.07 - 5.56) | 0.66 |
| **CXCL6** | | | | | | | | | | |
| Decrease | 20 (15 - 35) | 1 | | 1 | | 11 (8.7 - 14) | | | | |
| Increase | NR (NR - NR) | 0.24 (0.06 - 1.00) | 0.050 | 0.28 (0.06 - 1.40) | 0.12 | 14 (10 - NR) | 0.74 (0.26 - 2.08) | 0.56 | 0.59 (0.13 - 2.80) | 0.51 |
| **CXCL8** | | | | | | | | | | |
| Decrease | 29 (17 - NR) | 1 | | 1 | | 11 (9.8 - 17) | | | | |
| Increase | 20 (13 - NR) | 0.24 (0.06 - 1.00) | 0.050 | 1.25 (0.63 - 2.49) | 0.52 | 9.8 (7.0 - NR) | 0.74 (0.26 - 2.08) | 0.56 | 1.20 (0.52 - 2.74) | 0.67 |
| **CXCL12** | | | | | | | | | | |
| Decrease | 20 (15 - NR) | 1 | | 1 | | 11 (9.3 - 14) | | | | |
| Increase | NR (25 - NR) | 0.49 (0.15 - 1.60) | 0.24 | 0.81 (0.35 - 1.89) | 0.63 | 14 (9.8 - NR) | 1.03 (0.40 - 2.65) | 0.95 | 1.11 (0.63 - 1.96) | 0.71 |
| **CXCL9** | | | | | | | | | | |
| Decrease | 20 (14 - 35) | 1 | | 1 | | 11 (7.7 - 17) | | | | |
| Increase | NR (18 - NR) | 0.61 (0.31 - 1.21) | 0.16 | 0.65 (0.32 - 1.31) | 0.22 | 10 (8.9 - 20) | 0.85 (0.46 - 1.57) | 0.60 | 0.92 (0.50 - 1.69) | 0.79 |
| **CXCL10** | | | | | | | | | | |
| Decrease | 20 (14 - 32) | 1 | | 1 | | 11 (9.3 - 13) | | | | |
| Increase | 40 (16 - NR) | 0.62 (0.32 - 1.22) | 0.17 | 0.56 (0.27 - 1.19) | 0.13 | 10 (8.7 - NR) | 0.70 (0.37 - 1.32) | 0.27 | 0.65 (0.33 - 1.30) | 0.22 |
| **CXCL11** | | | | | | | | | | |
| Decrease | 30 (20 - NR) | 1 | | 1 | | 11 (9.8 - 20) | | | | |
| Increase | 16 (13 - NR) | 1.61 (0.82 - 3.16) | 0.17 | 1.52 (0.73 - 3.14) | 0.26 | 10 (7.5 - 17) | 1.46 (0.77 - 2.76) | 0.25 | 1.49 (0.76 - 2.93) | 0.25 |
| **CXCL13** | | | | | | | | | | |
| Decrease | 15 (13 - 32) | 1 | | 1 | | 8.9 (7.7 - 12) | | | | |
| Increase | 40 (25 - NR) | 0.37 (0.19 - 0.71) | 0.003 | 0.36 (0.15 - 0.85) | 0.019 | 14 (11 - 27) | 0.37 (0.19 - 0.71) | 0.003 | 0.32 (0.14 - 0.74) | 0.007 |
| **CXCL16** | | | | | | | | | | |
| Decrease | 27 (14 - NR) | 1 | | 1 | | 10 (7.8 - 14) | | | | |
| Increase | 23 (18 - NR) | 0.80 (0.42 - 1.56) | 0.52 | 0.73 (0.38 - 1.43) | 0.36 | 11 (9.8 - 27) | 0.64 (0.35 - 1.20) | 0.16 | 0.61 (0.33 - 1.11) | 0.11 |

Median OS and PFS were estimated with the Log Rank test. Hazard Ratios (HR), 95% Confidence Intervals (CI) and p-values, were obtained using the Cox proportional hazards models with or without the inverse probability weighting (IPW) approach. NR = Not reached

**Supplementary Table S6. Genes related to Tertiray Lymphoid Structures signatures (TLS)**

| TLS GENE SIGNATURES | | |
|---|---|---|
| **12-CHEMOKINE CELL SIGNATURE** | **TFH CELL SIGNATURE** | **TH1 AND B CELL SIGNATURE** |
| CCL2 | CXCL13 | CD4 |
| CCL3 | CD200 | CCR5 |
| CCL4 | FBLN7 | CXCR3 |
| CCL5 | ICOS | CSF2 |
| CCL8 | SGPP2 | IGSF6 |
| CCL18 | SH2D1A | IL2RA |
| CCL19 | TIGIT | CD38 |
| CCL21 | PDCD1 | CD40 |
| CXCL9 | | CD5 |
| CXCL10 | | MS4A1 |
| CXCL11 | | SDC1 |
| CXCL13 | | GFI1 |
| | | IL1R1 |
| | | IL1R2 |
| | | IL10 |
| | | CCL20 |
| | | IRF4 |
| | | TRAF6 |
| | | STAT5A |

THF = T-Follicular Helper cell; TH1 = T-helper cell
